# Regional and sex differences in the contribution of smoking-related mortality to changes in adult life expectancy in Brazil, 2000-23

**DOI:** 10.64898/2026.09.10.26362707

**Authors:** Julia Almeida Calazans, Fanny Janssen, Felipe Sanchez, José Manuel Aburto

## Abstract

**Background:** Since the 1980s, Brazil has implemented comprehensive tobacco control policies alongside major social changes, including improvements in education and reductions in poverty. These transformations contributed to a substantial decline in smoking prevalence and established Brazil as a global reference in tobacco control. However, the extent to which these changes have shaped recent mortality patterns and life expectancy across regions remains unknown. This study examines the contribution of smoking-related mortality to adult life expectancy in Brazil by sex and state from 2000 to 2023.

**Methods:** Smoking-related mortality was estimated using the Peto-Lopez method. Changes in life expectancy at age 35 were decomposed by age and cause of death using the linear integral method.

**Results:** Between 2000 and 2023, adult life expectancy at age 35 increased from 37.8 to 41.5 years among men and from 43.3 to 46.6 years among women. Among men, smoking-related mortality contributed 23.5% to this increase. States with the largest improvements, including Rio de Janeiro (36.4%) and Rio Grande do Sul (54.7%), showed the largest contributions from reductions in smoking-related mortality. Among women, increases in smoking-related mortality contributed negatively to changes in adult life expectancy (3.9%), driven by increased mortality after age 60. This negative contribution was observed in most states.

**Conclusion:** Trends in smoking-related mortality, partly driven by tobacco control policies, have contributed to increases in life expectancy among men, with important differences across states. Addressing sex and regional disparities in smoking mortality will support gains in life expectancy.

## INTRODUCTION

Smoking is a global public health challenge, and a major risk factor for chronic noncommunicable diseases such as cardiovascular conditions, cancer, and respiratory diseases.^1–5^ Despite being a global health concern, tobacco use disproportionately affects developing regions, with approximately 80% of users living in low- and middle-income countries^6^. In Brazil, smoking led to approximately 160,000 deaths in 2017, accounting for more than 12% of all deaths nationwide according to the Global Burden of Disease report.^7^

Since the 1980s, Brazil has implemented a broad set of public policies to control tobacco use under the National Tobacco Control Program, which were progressively expanded over time.^8.9^ Among the main measures are bans on advertising and sponsorship, smoke-free environments, pictorial health warnings on cigarette packages, tax increases, and minimum prices for tobacco products.^1,9–12^ These policies were implemented alongside broader social changes, including expansion in educational attainment and reductions in poverty, which contributed to a gradual decline in the social acceptability of smoking.^9^

As a result, smoking prevalence among adults declined from 32.1% in 2000 to 16.5% in 2018.^1,6^ These declines were observed across all states, both sexes, and all age groups.^2^ The magnitude of this reduction is particularly evident when compared with other Latin American countries, which in 2018 exhibited higher smoking prevalence, including Argentina (21.1%), Chile (43.0%), and Uruguay (20.4%).^6^ These achievements have established Brazil as an international benchmark in tobacco control.^10^

While several studies have examined smoking-related mortality in Brazil, they have not assessed the extent to which declines in smoking-related mortality have contributed to changes in adult life expectancy, or whether such contributions differ by sex and region. This is partly because reductions in smoking-related mortality typically occur two to three decades after declines in smoking prevalence begins,^11,12^ which makes their contribution to life expectancy a priori unknown. We expect changes in life expectancy to vary by sex and causes of death, supported by recent evidence reporting significant decreases in smoking-related mortality among adult men,^1,10,13^ with large reductions in mortality from cardiovascular diseases and chronic obstructive pulmonary diseases, and modest decreases in smoking-related cancers.^1,11^

We also hypothesise large regional variation given observed trends in smoking-related mortality recently: large declines in the most developed in South and Southeast, that historically high tobacco-related mortality. In contrast, reductions have been limited in less developed states, particularly those in the North and Northeast.^1^

Understanding how these trends in the past couple of decades have translated into adult life expectancy changes, and their distribution across the country, remains an open question with important implications for both tobacco control policies and future life expectancy advances. In this context, this study examines the contribution of smoking-related mortality to adult life expectancy and its changes over time in Brazil by sex and state from 2000 to 2023.

## METHODS

### Mortality data

We use two complementary data sources to estimate age- and cause-specific mortality rates. All-cause age-specific mortality rates were obtained from the Brazilian Institute of Geography and Statistics (IBGE) from 2000 to 2023. These rates are adjusted for underreporting and other quality issues in death registration. Cause-of-death microdata were obtained from the Mortality Information System (SIM) of the Brazilian Ministry of Health, which include sex, age, state of residence, and underlying cause of death. Missing age or sex accounted for approximately 0.3% of all deaths and was redistributed proportionally based on the distribution of deaths with complete information. Age- and cause-specific mortality rates were then calculated as the product of the cause-specific proportion of deaths from SIM and the corresponding all-cause mortality rate from IBGE. The list of states and their respective regions is provided in Supplementary Material A.

### Smoking-Related Deaths Definition

Smoking-related mortality among adults aged 35 years and older was estimated using the indirect Peto-Lopez method,^15^ which has been widely validated and applied across multiple countries, including Brazil. ^10,16^ This method uses lung cancer mortality as a biomarker of cumulative population-level tobacco exposure. Smoking-related mortality from other causes is then derived by combining this exposure measure with cause-, sex-, and age-specific relative risks for smokers and non-smokers from the American Cancer Society’s Cancer Prevention Study II (Table 1B and 2B in the Supplementary Material B).

Smoking-related mortality was estimated by aggregating deaths across six cause groups: 1) smoking-related lung cancer; 2) other smoking-related cancers (including upper aerodigestive cancers); 3) smoking-related chronic respiratory diseases; 4) infectious respiratory diseases (tuberculosis, pneumonia, and influenza); 5) smoking-related cardiovascular diseases; and 6) other smoking-related causes. Five non-smoking-related cause groups were also analyzed: 1) non-smoking-related communicable causes (infectious diseases, maternal, perinatal, and congenital causes); 2) external causes; 3) alcoholic liver cirrhosis; 4) non-smoking-related cardiovascular diseases; and 5) other non-smoking-related causes. ICD codes for all groups are detailed in Supplementary Material C.

In addition to age-specific mortality rates, we estimated age-standardized smoking-related mortality trends over time using the average population of men and women in 2000 as the standard population.

### Cause-deleted life tables

The burden of smoking-related mortality on life expectancy at age 35 in 2000, 2019 and 2023 was estimated as the potential gain in life expectancy (PGLE) with cause-deleted life tables, assuming that mortality from a certain cause is hypothetically set to zero while all other causes remain contributors to mortality. Results are expressed as relative gains in life expectancy at age 35, enabling comparisons across states and over time. Absolute gains are presented in the Supplementary Material D.

### Demographic decomposition

The contribution of smoking-related mortality to changes in adult life expectancy over time (2000-2023) was estimated using the linear integral decomposition method.^17^ This approach quantifies the contribution of each age group and cause of death to differences in life expectancy at age 35 between two periods, allowing the impact of smoking-related mortality to be distinguished from that of other causes.

### Sensitivity analysis

The results for 2000-2023 were compared with the results for the pre-pandemic period (2000–2019) to assess the influence of the COVID-19 pandemic.

### Uncertainty analysis

Confidence intervals (95% CI) for number of deaths, mortality rates, and potential gains in life expectancy were estimated using Monte Carlo simulation with a zero-truncated normal distribution, while a normal distribution was used for decomposition contributions.

### Replicability

All analyses were performed in R version 4.5.1. Complete code for data processing, statistical analysis, and visualization is publicly available at [GitHub link].

## RESULTS

### Trends in life expectancy at age 35

Between 2000-2023, life expectancy at age 35 increased from 37.8 to 41.5 years for men and from 43.3 to 46.6 years for women. This progress has been uneven across states (Figure 1). Overall, there was evidence of regional convergence, with lowest life expectancy states experiencing the largest improvements between 2000-2023. Most notably in Sergipe and Tocantins, where female life expectancy increased by more than five years over time, and in Minas Gerais, Santa Catarina, Rondônia, and Rio de Janeiro for men, with increases of more than four years. In 2023, the lowest female life expectancy at age 35 was observed in Alagoas and Rio de Janeiro while the highest levels were found in Piaui and the Distrito Federal. Among men, the lowest life expectancy at age 35 was observed in Alagoas and Pernambuco, while the highest levels were found in Santa Catarina, and the Distrito Federal.

**Figure 1.**
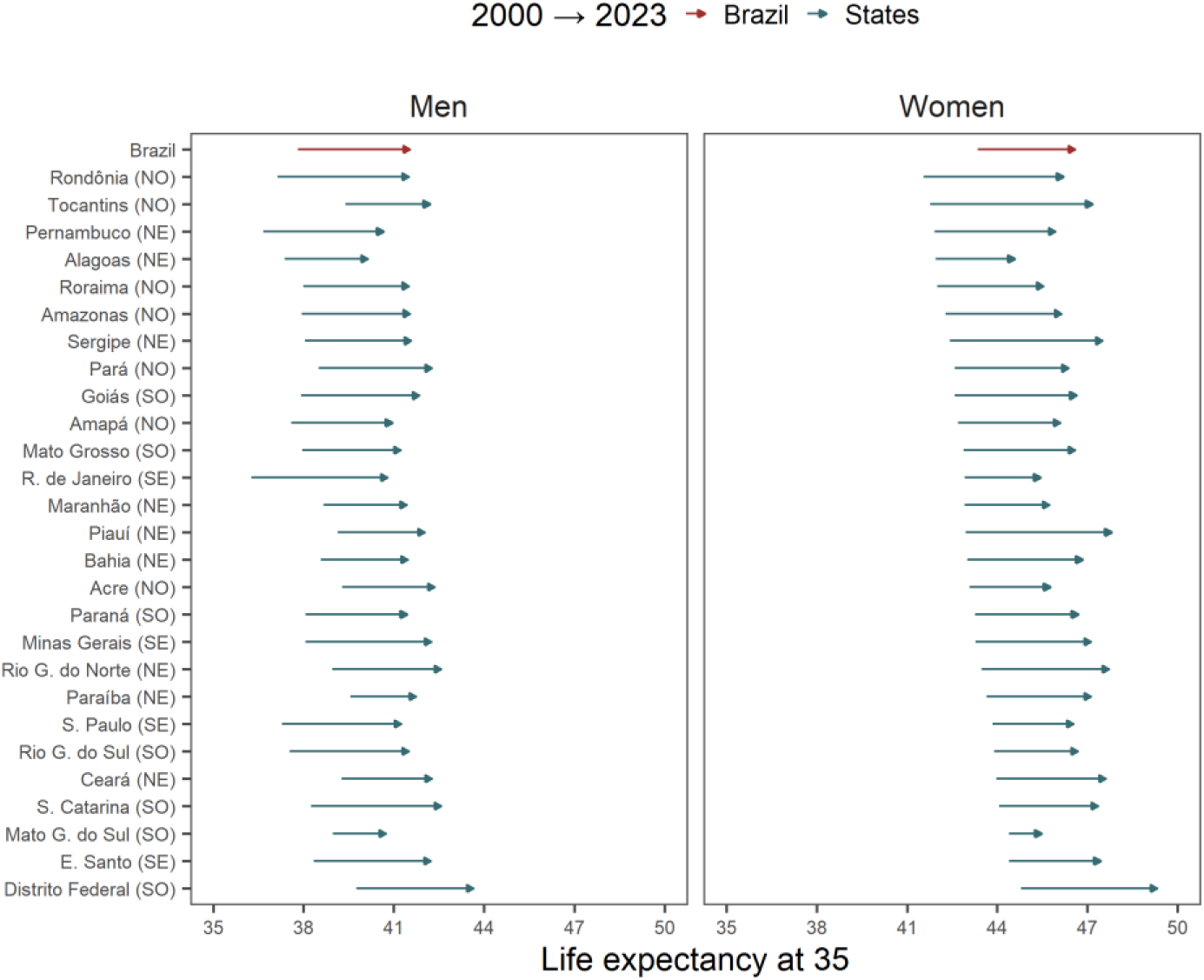
Life expectancy at age 35 – Men and Women, Brazilian states (2000 and 2023) Note: States are ordered according to female life expectancy at age 35 in 2000. NO = North, NE = Northeast, CW = Central-West, SE = Southeast, SO = South. **Source:** SIM/DATASUS/MS and IBGE.

Adult life expectancy in 2023 exceeded 2019 levels in Brazil, indicating recovery of the pre-pandemic upward trend, although some states, particularly in the North and Northeast, had not yet fully recovered their pre-pandemic trajectories by 2023 (Tables D2 and D3 in Supplementary Material).

### Trends in smoking-related mortality rates

Between 2000 and 2023, age-specific smoking-related mortality rates declined substantially among men (Figure 2), particularly at ages 50–79. Among individuals aged 85 years and older, trends were less consistent, with rates increasing over time. In contrast, women experienced modest declines in smoking-related mortality at ages 35–59, followed by a progressively steeper rise in mortality from age 60 onwards. Women aged 85 years and older show the largest increases in smoking-related mortality over time. These trends are reflected in age-standardized smoking-related mortality rates (Tables D1 and D2 in Supplementary Material). In 2000, age-standardized smoking-related mortality was 19.37 per 10,000 for men and 5.55 for women. By 2023, the male rate had fallen to 9.44 per 10,000, while the female rate increased slightly to 6.64 per 10,000.

**Figure 2.**
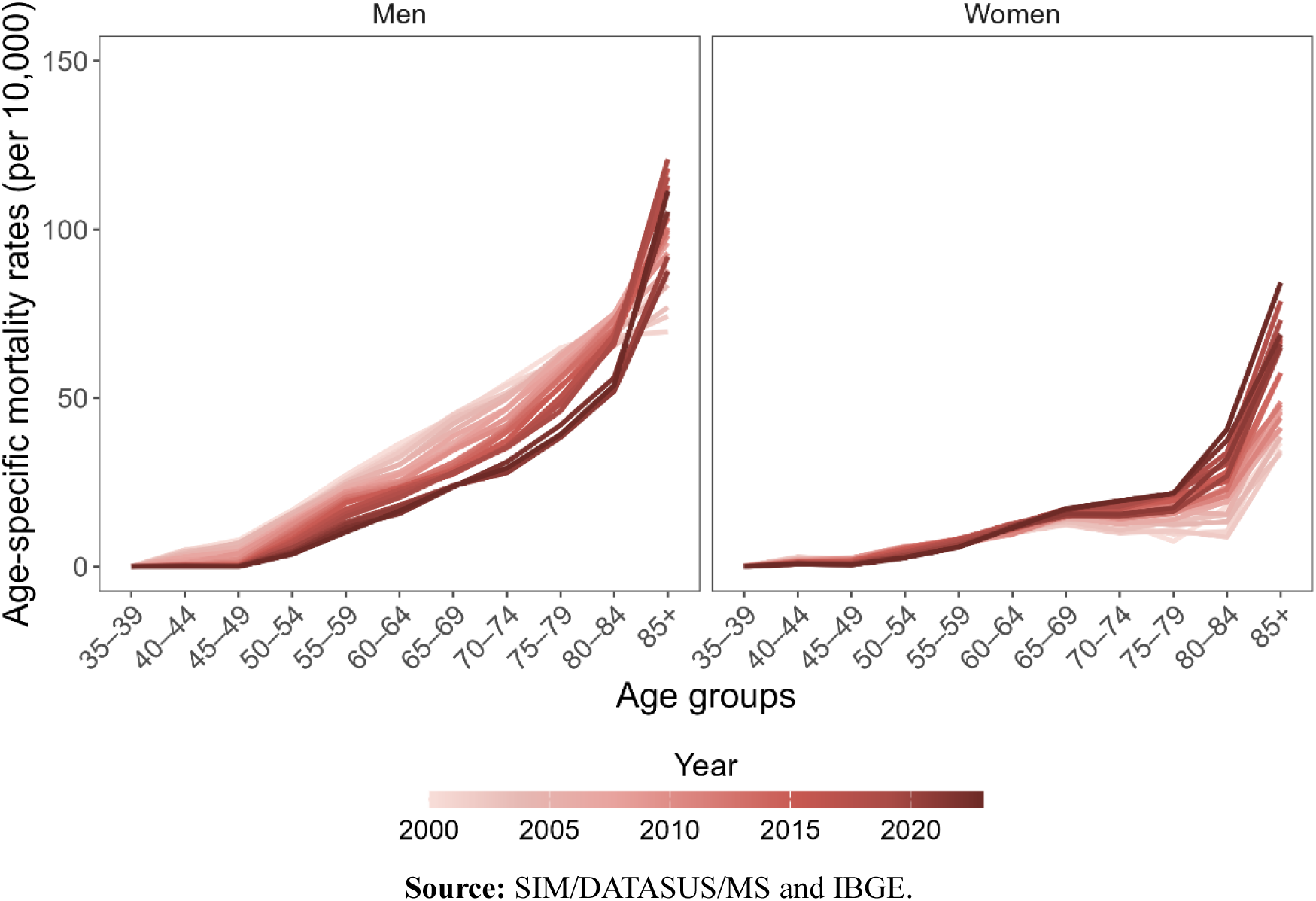
Age-specific smoking-related mortality rates – Men and women, Brazil (2000 to 2023) Source: SIM/DATASUS/MS and IBGE.

### The burden of smoking-related mortality on life expectancy at age 35

In 2000, the burden of smoking-related mortality on life expectancy at age 35, measured as the potential gain derived from the theoretical elimination of smoking-related mortality (PGLE), was 1.45 years (4.05%) among men and 0.6 years (1.37%) among women. By 2023, this burden had declined to 0.89 years (2.15%) among men while increasing slightly to 0.9 years (1.93%) among women (Figure 3 and Tables D3 and D4 in Supplementary Material).

**Figure 3.**
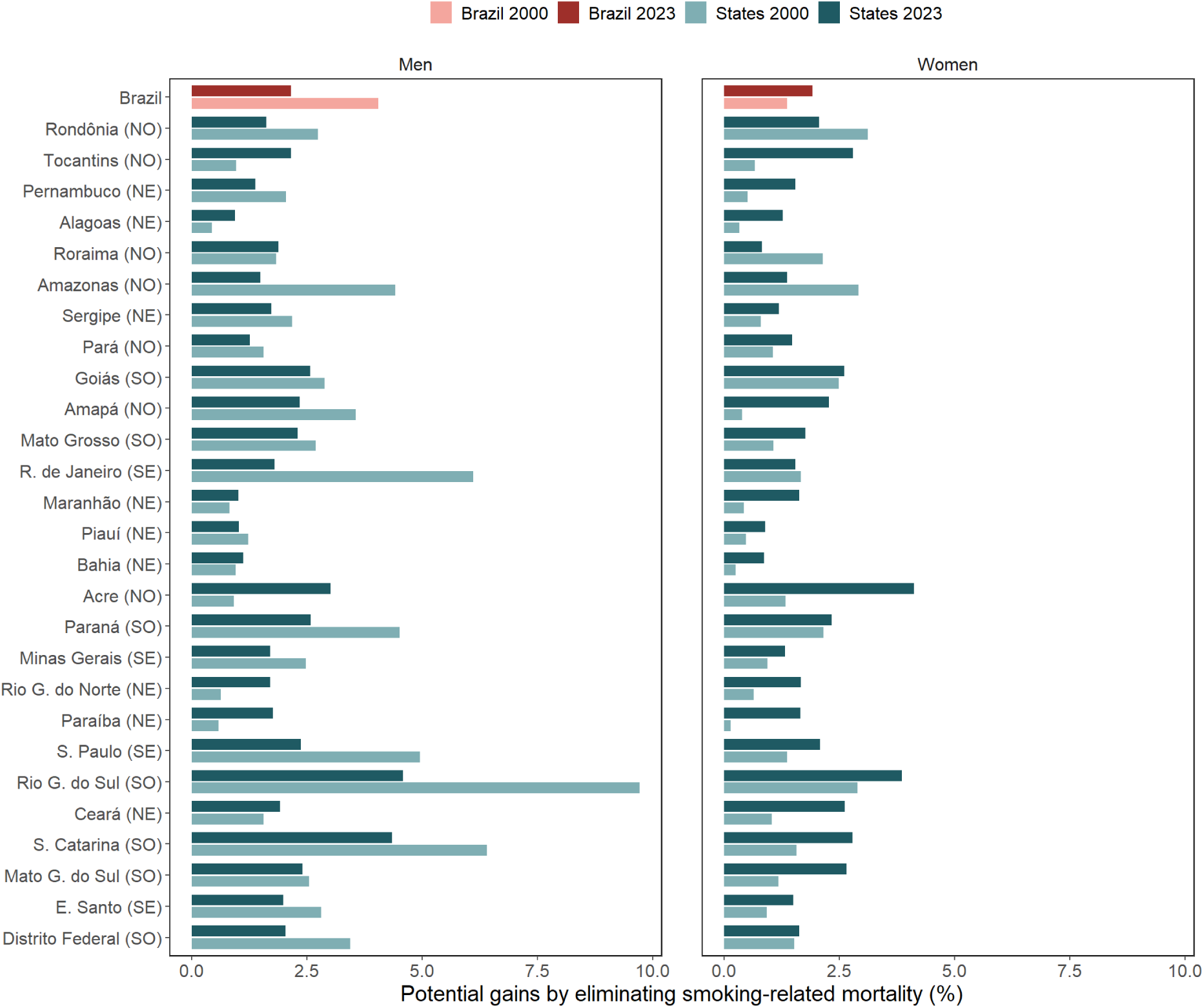
Potential gain in life expectancy at 35 by eliminating smoking-related mortality – Men and women, Brazilian states (2000 and 2023) Note: States are ordered according to female life expectancy at age 35 in 2000. NO = North, NE = Northeast, CW = Central-West, SE = Southeast, SO = South. **Source:** SIM/DATASUS/MS and IBGE.

In 2000, the burden was highest among women in Rondônia, Amazonas, Rio Grande do Sul, and Goiás, and lowest in the North and Northeast, particularly in Amapá, Alagoas, Bahia, and Paraíba. Between 2000-2023, the burden raised in most states, with the largest improvements in Tocantins and Acre, and declined in Rio de Janeiro, Rondônia, Roraima, and Amazonas. Among men, the burden was highest in the South and Southeast, especially in Paraná, São Paulo, Rio de Janeiro, Santa Catarina, and Rio Grande do Sul, where smoking-related mortality reduced life expectancy by more than 4.5 years, and lowest in the North and Northeast. This pattern persisted in 2023, although the burden declined in most states, with exceptions in several North and Northeast states (Roraima, Bahia, Maranhão, Ceará, Alagoas, Rio Grande do Norte, Paraíba, Tocantins, and Acre) and the largest declines in Rio Grande do Sul and Rio de Janeiro.

Overall, trends were consistent in 2000, 2019, and 2023 (Tables D3 and D4 in Supplementary Material), with the burden increasing among women and declining among men nationally. Most states followed the same pattern, apart from Roraima and Goiás among men, and Rondônia, Pará, Rio de Janeiro, and Mato Grosso among women.

### Contribution of smoking-related mortality to changes over time in life expectancy at age 35

Between 2000-2023, life expectancy at age 35 increased by 3.7 years among men in Brazil, of which 0.88 year (23.5%) was attributable to reductions in smoking-related mortality (Figure 4). This contribution was larger than that observed for the 2000–2019 period (Table D5 in Supplementary Material). Among smoking-related causes, cardiovascular diseases contributed the most (6.9% of the total increase), followed by lung and other smoking-related cancers combined (6.8%). All non-smoking-related causes contributed positively, particularly cardiovascular diseases (34.6%) and other causes (35.7%).

**Figure 4.**
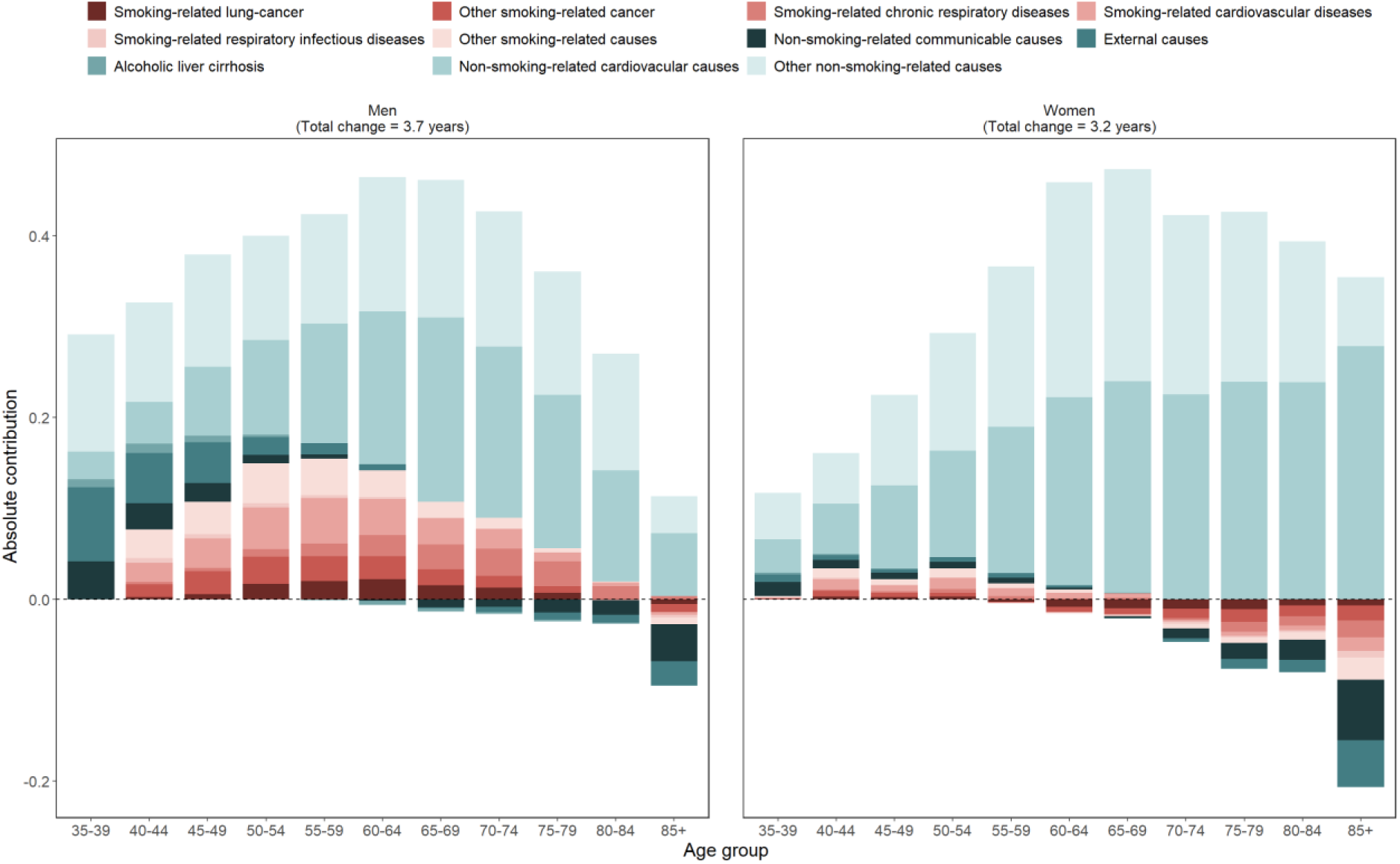
Decomposition of the change in life expectancy at age 35 by cause of death and age group – Men and women, Brazil (2000 to 2023) Note: Smoking-related causes are shown in red shades, and non-smoking-related causes are shown in green shades. Source: SIM/DATASUS/MS and IBGE.

The contribution of smoking-related causes raised progressively from ages 35 to 59 and declined thereafter. At ages 85 and older, smoking-related mortality growth over the period, reducing life expectancy at age 35 by 0.03 year (0.68%), driven primarily by smoking-related cancers. Among non-smoking-related causes, infectious diseases and external causes also had a negative effect, while all remaining causes contributed positively.

Among women, life expectancy at age 35 increased by 3.2 years between 2000-2023, but changes in smoking-related mortality reduced this improvement by 0.13 year (3.9%) (Figure 4). The negative contribution of smoking-related mortality was slightly smaller, although not statistically significant, for the 2000–2019 period (Table D6 in Supplementary Material). All smoking-related causes had a net negative effect except cardiovascular diseases, which contributed positively (0.9%). Cancers had the largest negative effect (2.9%). Among non-smoking-related causes, cardiovascular diseases (34.6%) and other causes (35.7%) were the largest positive contributors.

Regarding the age pattern, smoking-related causes had a positive effect up to age 59, with ages 40–54 contributing the most (2.5%). From 60 onward, contributions became systematically negative, particularly at ages 75–84 (−2.5%). This pattern varied by cause: cancers and infectious respiratory diseases showed negative contributions from age 55 onward, chronic respiratory diseases from age 70, and other causes from age 65.

### Regional disparities in the contribution of smoking-related mortality to changes in life expectancy at age 35

Among men, reductions in smoking-related mortality contributed positively to life expectancy improvement in most states (Figure 5). Exceptions were eight states in the North and Northeast: Acre (−23.5%), Paraíba (−22.2%), Rio Grande do Norte (−11.2%), Tocantins (−10.2%), Alagoas (−7.1%), Ceará (−2.1%), Roraima (−2.0%), and Maranhão (−2.0%), where negative effects were driven primarily by other smoking-related cancers. The largest positive contributions were observed in Rio Grande do Sul (54.7%), Rio de Janeiro (36.4%), São Paulo (30.7%), Amazonas (29.7%), Santa Catarina (27.7%), and Paraná (25.9%), where reductions in smoking-related cardiovascular mortality were the main driver. In states with negative contributions, such as Acre, Alagoas, Paraíba, and Tocantins, negative effects extended across nearly the entire age distribution, whereas states with positive contributions mirrored the national age pattern (Figure 1D in Supplementary Material).

**Figure 5.**
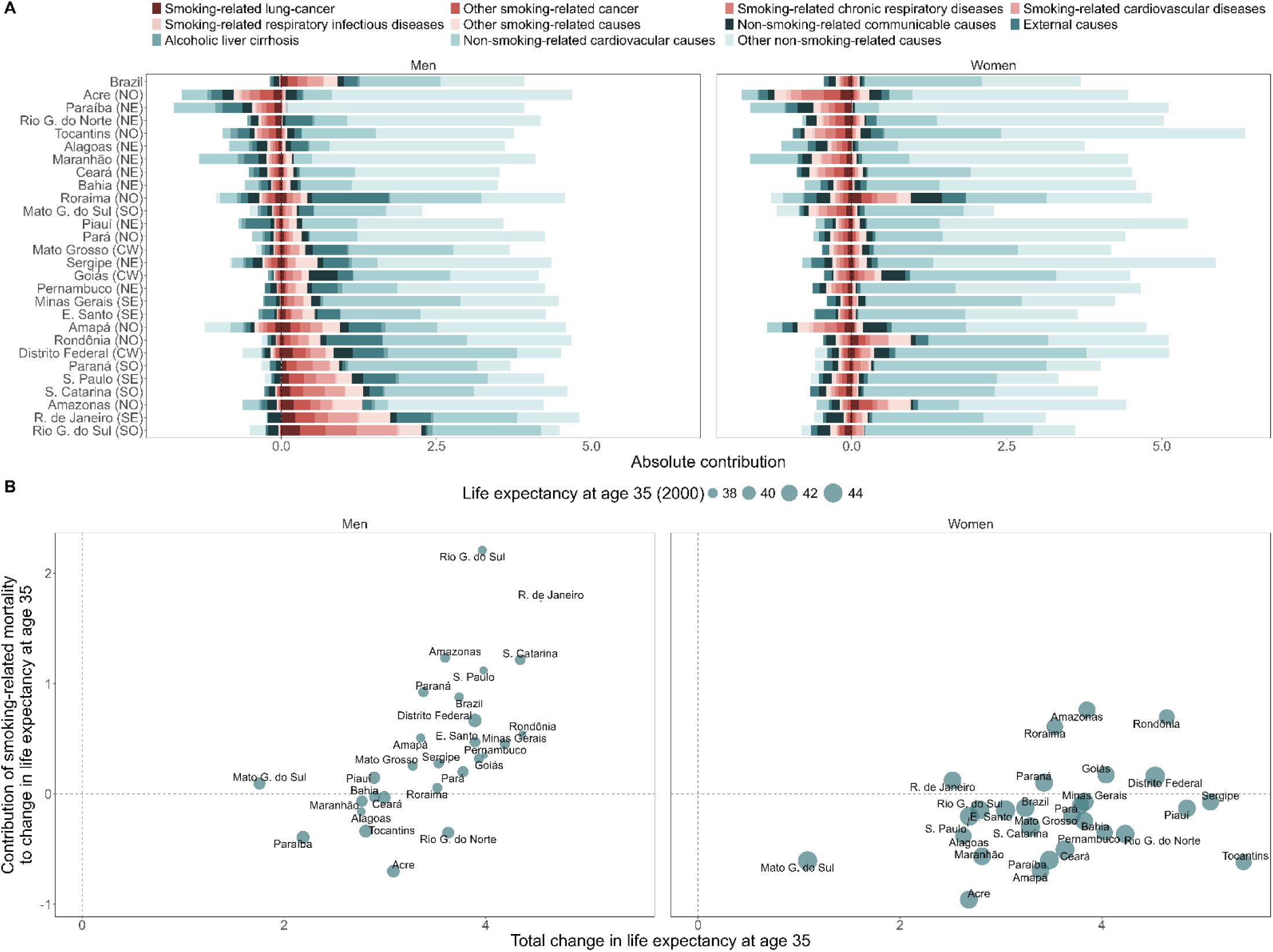
Contributions of smoking-related mortality to changes in life expectancy at age 35 – Men and Women, Brazilian states (2000–2023) Note: A) Decomposition of changes in life expectancy at age 35 by cause of death, Brazilian states (2000–2023). B) Contribution of smoking-related mortality to changes in life expectancy at age 35 versus total life expectancy gain, Brazilian states (2000–2023). In the panel A, the states are ordered according to male smoking-related contribution to changes in life expectancy at age 35. Smoking-related causes are shown in red shades, and non-smoking-related causes are shown in green shades. NO = North, NE = Northeast, CW = Central-West, SE = Southeast, SO = South. **Source:** SIM/DATASUS/MS and IBGE.

Among women, changes in smoking-related mortality contributed negatively to life expectancy in most states (Figure 4), with positive contributions observed only in Amazonas (23.3%), Rondônia (16.1%), Roraima (15.0%), Goiás (14.6%), the Distrito Federal (5.9%), and Paraná (2.5%). The largest negative contributions were in Mato Grosso do Sul (−42.6%), Acre (−28.5%), Maranhão (−22.7%), and Paraíba (−22.3%). In states with negative contributions, the age pattern resembled the national level, while in states with positive contributions, such as Amazonas, Roraima, and Rondônia, no clearly defined age pattern was observed (Figure 2D in Supplementary Material). Results for the 2000–2019 period follow the same pattern as those observed for 2000–2023 for both sexes.

States with larger total improvements in life expectancy at age 35 tended to show greater positive contributions from reductions in smoking-related mortality (Figure 5), and this association was more pronounced among men than among women. Among men, Rio de Janeiro and Rio Grande do Sul stand out as outliers, with particularly high values for both total life expectancy improvements and the contribution of smoking-related mortality. Among women, the association was weaker, reflecting more heterogeneous trends across states.

## DISCUSSION

This is first study to comprehensively assess the contribution of smoking-related mortality to changes in adult life expectancy in Brazil by sex and state, contributing to existing literature on how reductions in smoking have driven recent mortality declines, particularly from cardiovascular and chronic diseases.^18^ Our findings reveal divergent trends between men and women. Among men, the burden of smoking-related mortality on life expectancy at age 35 declined from 4.05% to 2.15% between 2000-2023, with reductions in smoking-related mortality contributing to 23.5% of the total increase in adult life expectancy. Among women, a slight increase in smoking-related mortality, particularly from age 60 onward, reduced life expectancy at age 35 by 3.9%, increasing the burden from 1.37% to 1.93%. These trends are consistent with Palloni and colleagues,^19^ who documented increasing impacts of smoking on female adult life expectancy across Latin America between 1980-2009.

The trends in smoking-related mortality in Brazil are consistent with the smoking epidemic model described by Lopez et al.,^11^ which characterizes a four-stage transition in smoking prevalence and smoking-related mortality with important differences in timing and level between sexes. Women took up smoking later than men, and their smoking-related mortality therefore peaks later and at lower levels. As documented by Janssen^20^ for low-mortality countries, the timing of the maximum mortality impact differs between sexes by more than 25 years on average. As a result, the simultaneous decline in smoking-related mortality among men and increase among women is consistent with our findings. Similar patterns have also been observed in some European countries, where, although smoking-related mortality among women remains substantially lower than among men, the gap has narrowed markedly over time.^20–23^

The increase in smoking-related mortality among women can be largely explained by cohort differences in social context. Women born before 1940, who were aged 60 or older in 2000, reached adulthood when smoking among women was uncommon and strongly socially stigmatized. In contrast, cohorts born in subsequent decades reached early adulthood in the 1970s and 1980s, when greater female participation in education and the labor market coincided with tobacco industry campaigns associating smoking with modernity and emancipation, marking the peak of smoking diffusion among women.^12,14,24,25^ Smoking prevalence was approximately 30.7% among women born between 1934–1943, rising to 47.7% among those born between 1954–1963.^12^

These findings do not imply that tobacco control policies have been ineffective among women. In fact, smoking-related mortality declined among women aged 35–59 between 2000-2023. However, the effects of past smoking exposure among older female cohorts continue to shape current mortality patterns, and reductions at younger ages have not yet been sufficient to offset the higher burden among older women with historically higher smoking prevalence. There is therefore an urgent need for policies aimed at reducing the health and mortality consequences of smoking, particularly among older female cohorts.

Our findings reveal marked regional inequalities in the contribution of smoking-related mortality to changes in life expectancy at age 35, confirming our initial hypothesis. The states where smoking-related mortality contributed negatively are concentrated in the North and Northeast, while most states in the South, Southeast, and the Federal District showed positive contributions, particularly among men. Furthermore, the states with the greatest overall advances in life expectancy at age 35 tended to show greater positive contributions from the reduction in smoking-related mortality, especially Rio de Janeiro and Rio Grande do Sul. This pattern suggests that regional dynamics depend not only on historical smoking prevalence but also on structural conditions and health system responsiveness.^1^ South and Southeast have historically exhibited high smoking prevalence, also concentrate higher socioeconomic development, which may have facilitated more effective policy implementation and more timely management of smoking-related diseases. In contrast, larger socioeconomic vulnerability and health system challenges in the North and Northeast may explain why smoking impacts translate more persistently into longevity losses, particularly among women.

### Limitations

This study has several limitations. First, part of the regional differences in all-causes mortality may reflect assumptions in estimating life tables, which are projected based on census years and may not accurately capture annual mortality risks. Nevertheless, using official statistics allows greater comparability with other studies. Second, cause-specific mortality rates may show artificial increases due to improvements in death registration quality rather than true rises in mortality risk, particularly in the North and Northeast, where registration quality has historically been more limited. On the other hand, evidence indicates that the quality of mortality information has improved substantially since the 2000s, allowing more appropriate comparisons across regions.^26,27^ Third, wide confidence intervals in some Northern states reflect the small number of smoking-related deaths in these low-population areas. Fourth, the use of relative risk parameters from the U.S. Cancer Prevention Study II may not fully capture differences in smoking-related mortality across populations and over time.^28^ More importantly, the same sex-specific relative risks were applied uniformly across all states, which does not account for potential regional variation in smoking-related mortality risks. However, this limitation can be less consequential for the analysis of trends over time. Alternative methods, such as that proposed by Preston and colleagues,^29^ rely on data from countries with substantially higher smoking prevalence than Brazil, which could further distort estimates. Fifth, the study considers only smoked tobacco products, excluding smokeless tobacco and electronic cigarettes. However, the prevalence of these products in Brazil remains below 0.3%,^1^ suggesting a limited impact on aggregate results.

This study is limited to mortality from age 35 onward, in line with the Peto–Lopez method, which captures the long-term cumulative effects of smoking on mortality. However, the recent upsurge in electronic cigarette use and other nicotine products among young people is a cause for concern.^30^ Although its effects on adult mortality cannot yet be measured, this pattern may represent an important public health challenge in the coming decades, particularly given the potential cardiovascular and other long-term consequences of early nicotine exposure.^31^

Finally, this study does not directly assess the impact of tobacco control policies on adult life expectancy in Brazil; rather, it examines the contribution of changes in smoking-related mortality to life expectancy in a declining tobacco use context. While tobacco control policies may have contributed to this reduction, other concurrent social, economic, and cultural changes may also have influenced declines in tobacco use and related mortality.

### Public Health Implications

Tobacco use is a complex issue with social, cultural, economic, and public health dimensions.^32,33^ This study indicates that tobacco control policies implemented in Brazil since the 1980s have been associated with substantial reductions in smoking-related mortality among men. However, effects among women have been more limited, highlighting the need for targeted strategies for smoking prevention and control among women, as well as policies to mitigate long-term health consequences, particularly among older populations. Continuous monitoring of smoking indicators is essential to track progress toward national and international targets, assess tobacco control policies, and guide future interventions. Regional disparities in both smoking prevalence and smoking-related mortality further suggest that national policies should be complemented by context-specific interventions that consider socioeconomic vulnerability, health system organization, and historical patterns of tobacco use. These patterns have direct implications for future mortality trends, as changes in smoking-related mortality are likely to shape trajectories of adult life expectancy in the coming decades. Beyond tobacco control, these findings also have implications for policies aimed at reducing inequalities in life expectancy, particularly across regions and between sexes.

## Data Availability

All analyses were performed in R version 4.5.1. Complete code for data processing, statistical analysis, and visualization is publicly available at [The GitHub link will be provided after the peer review process to preserve anonymity during peer review].

## AUTHOR CONTRIBUTIONS

JAC and JMA conceptualized and designed the study. JAC carried out the analysis and produced results and FS, JMA and FJ contributed to the interpretation of the results. JAC wrote the first draft of the paper. All authors commented on previous versions of the manuscript, as well wrote and approved the final manuscript.

## DECLARATION OF GENERATIVE AI AND AI-ASSISTED TECHNOLOGIES

ChatGPT (OpenAI, GPT-5.5 mini) was used solely to improve the clarity, readability, and English language of the manuscript. All content was reviewed, edited, and approved by the authors, who take full responsibility for the accuracy and integrity of the work.

## SUPPLEMENTAL MATERIAL

### Supplemental Material A - List of Brazilian states by region

| Regions | States |
| --- | --- |
| North (NO)<br>(7 states) | Acre (AC), Amapá (AP), Amazonas (AM), Pará (PA), Rondônia (RO), Roraima (RR), Tocantins (TO). |
| Northeast (NE)<br>(9 states) | Alagoas (AL), Bahia (BA), Ceará (CE), Maranhão (MA), Paraíba (PB), Pernambuco (PE), Piauí (PI), Rio Grande do Norte (RN), Sergipe (SE). |
| Central-West (CW)<br>(3 states + DF) | Goiás (GO), Mato Grosso (MT), Mato Grosso do Sul (MS), Distrito Federal (DF). |
| Southeast (SE)<br>(4 states) | Espírito Santo (ES), Minas Gerais (MG), Rio de Janeiro (RJ), São Paulo (SP). |
| South (SO)<br>(3 states) | Paraná (PR), Rio Grande do Sul (RS), Santa Catarina (SC). |

### Supplemental Material B - Detailed Description of the Peto-Lopez Method

The Peto–Lopez method (Peto et. al. 1992) was employed to estimate the smoking related mortality rates from 2000 to 2023 in the Brazilian states. Conceptually, the method uses excess lung cancer mortality as an indicator of the cumulative hazards of smoking in both the study population and the reference population from the American Cancer Society Cancer Prevention Study II (CPS-II). This approach allows the accumulated impact of smoking in the population to be inferred indirectly from lung cancer mortality, which is strongly associated with long-term tobacco exposure.

### Calculation of Lung Cancer Mortality Rates

Age-specific lung cancer mortality rates by sex and state in Brazil were calculated as the product of the proportion of deaths attributed to lung cancer in death records and the all-cause mortality rates obtained from the Brazilian Institute of Geography and Statistics (IBGE) life tables.

### Lung Cancer Mortality Rates among Smokers and Never Smokers (CPS-II)

Table B1 presents the age-specific lung cancer mortality rates among smokers and never smokers in the reference population from the American Cancer Society Cancer Prevention Study II (CPS-II). Following the recommendations of Jansen et al. (2021), these age-specific mortality rates were smoothed separately for men and women and for smokers and never smokers to reduce random fluctuations across age groups.

**Table B1.** Annual lung cancer mortality rates (per 100,000) from years 3 to 6 inclusive (approximately 198<u>4-88) of ACS CPS-II prospective study of one million US ad</u>ults.

| Age group | Men |  | Women |  |
| --- | --- | --- | --- | --- |
|  | lung cancer<br>(smoker) | lung cancer<br>(non-smoker) | lung cancer<br>(smoker) | lung cancer<br>(non-smoker) |
|  | *C* | *A* | *C* | *A* |
| 35 | 0 | 7 | 7 | 3 |
| 40 | 23 | 0 | 12 | 0 |
| 45 | 35 | 9 | 49 | 4 |
| 50 | 114 | 5 | 71 | 5 |
| 55 | 227 | 3 | 136 | 8 |
| 60 | 375 | 11 | 195 | 14 |
| 65 | 599 | 24 | 310 | 18 |
| 70 | 899 | 36 | 339 | 28 |
| 75 | 1168 | 38 | 429 | 42 |
| 80+ | 1191 | 88 | 400 | 64 |

### Calculation of the Smoking Impact Ratio (SIR)

The Smoking Impact Ratio (SIR) measures the cumulative exposure to smoking in the study population relative to a reference population from the American Cancer Society Cancer Prevention Study II (CPS-II). It is calculated as:

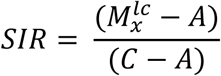

where 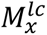 is the age-specific lung cancer mortality rate in the observed population; *A* is the age-specific lung cancer mortality rate among never smokers in the CPS-II reference population; and *C* is the age-specific lung cancer mortality rate among smokers in the CPS-II reference population.

Higher values of the SIR indicate greater cumulative exposure to smoking in the study population and, consequently, a larger share of deaths expected to be attributable to tobacco use. A value of SIR equal to 1 indicates that lung cancer mortality in the observed population is equivalent to that of smokers in the reference population, suggesting a similar accumulated exposure to smoking. Conversely, a value of SIR equal to 0 indicates that lung cancer mortality in the observed population is equal to that observed among never smokers in the reference population, implying no excess mortality attributable to smoking. When 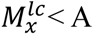< A, the SIR is set to 0.

### Application of Relative Risks

Relative risks of death (Table B2) measure the excess mortality associated with smoking for specific causes of death. These estimates were obtained from the American Cancer Society Cancer Prevention Study II (CPS-II) prospective study, as proposed by Peto et al. (1992). In addition, we followed the approach suggested by Ezzati and Lopez (2003), in which external causes, alcoholic liver cirrhosis, infectious diseases (except tuberculosis, pneumonia, and influenza), as well as maternal, perinatal, and congenital conditions, are not attributed to smoking.

**Table B2.** Cigarette smokers versus “non-smokers” (never smoked regularly). Selected risk ratios from years 3 to 6 inclusive (approximately 1984-88) of ACS CPS-II prospective study of one million us adults.

| Cause (ICD-9) | Male | Female |
| --- | --- | --- |
| Lung cancer (ICD 162) | 24.22 | 12.50 |
| Upper aerodigestive cancer – mouth, pharynx, larynx or oesophagus (ICD 140-150 and 161) | 7.87 | 6.95 |
| Other cancer (rest of ICD 140-209) | 1.69 | 1.20 |
| Chronic obstructive pulmonary disease (ICD 490-2, 492-6) | 13.82 | 14.21 |
| Other medical causes |  |  |
| Ages 35-59 | 3.05 | 2.69 |
| “ 60-64 | 2.31 | 2.68 |
| “ 65-69 | 2.09 | 2.52 |
| “ 70-74 | 2.00 | 2.00 |
| “ 75+ | 1.54 | 1.44 |

### Proportion of Deaths Attributable to Smoking (SAF)

For lung cancer, the proportion of deaths attributable to smoking follows the approach proposed by Gorini et al. (2003) and is defined as:

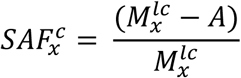

where 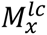 is the age-specific lung cancer mortality rate in the observed population and *A* is the lung cancer mortality rate among never smokers in the reference population from the American Cancer Society Cancer Prevention Study II (CPS-II). In this formulation, the smoking-attributable fraction corresponds to the excess lung cancer mortality relative to the total observed lung cancer mortality rate.

For other causes of death, the smoking-attributable fraction incorporates both the Smoking Impact Ratio (SIR) and the relative risks of mortality:

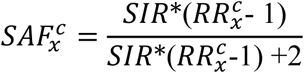

where *RR* 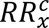 represents the relative risk of death for cause *c* in age group *x*.

### Estimation of Smoking- Related Deaths

For each age group and cause of death, the number of deaths attributable to smoking is estimated by multiplying the total number of deaths from that cause by the corresponding smoking-related fraction (SAF). The number of smoking- related deaths is first calculated for each state. National estimates for Brazil are then obtained by summing the state-level results, ensuring consistency between subnational and national totals.

### Estimation of Smoking-Related Mortality Rates

Finally, smoking-related mortality rates were estimated by applying the smoking- related fractions to the cause- and age-specific mortality rates obtained from the Brazilian Institute of Geography and Statistics (IBGE) life tables. Total smoking-related mortality rates were calculated by summing cause-specific smoking-related mortality rates across all smoking-related cause groups.

---

Supplemental Material C Classification of Smoking-Related and Non-Smoking-Related Causes (ICD-10)

| **Cause** | **ICD-10** |
| --- | --- |
| ***Smoking-related causes*** |  |
| Lung cancer | C33–C34 |
| Other cancer | Upper aerodigestive cancer (C00–C16, C18–C21, C22, C25, C32, C53, C64–C68)<br>Other cancers (C, D0-D4) |
| Chronic respiratory diseases | J40–J44, J47 |
| Cardiovascular diseases | Vascular diseases (I20-I25, I60)<br>Other cardiovascular diseases (I) |
| Infectious respiratory diseases | Tuberculosis (A15-A19)<br>Pneumonia and influenza (J00-J22) |
| Other causes | Other medical causes associated with smoking. |
| ***Non-smoking-related causes*** |  |
| Communicable causes | Infectious diseases (A-B, except A15-A19)<br>Maternal, perinatal, and congenital causes (O00–O99, P00–P96, Q00–Q99) |
| External causes | V01–Y89 |
| Alcoholic liver cirrhosis | K703 |
| Non-related cardiovascular diseases | The residual effect of cardiovascular causes (I) not associated with smoking. |
| Other non-related causes | All other causes |

## Supplemental Material D - Figures and Tables

**Tables D1.** Potential gain in life expectancy at age 35 (PGLE35) by eliminating smoking-related mortality – Men, Brazilian states (2000, 2019 and

|  | 2000 |  |  | 2019 |  |  | 2023 |  |  |
| --- | --- | --- | --- | --- | --- | --- | --- | --- | --- |
|  | Life expectancy at 35 | Absolute PGLE35 | Percentual PGLE35 | Life expectancy at 35 | Absolute PGLE35 | Percentual PGLE35 | Life expectancy at 35 | Absolute PGLE35 | Percentual PGLE35 |
| **Brazil** | **37.81** | **1.53** | **4.05** | **41.32** | **1.07** | **2.6** | **41.55** | **0.89** | **2.15** |
|  |  | **(1.49,1.58)** | **(3.93,4.17)** |  | **(1.05,1.10)** | **(2.53,2.66)** |  | **(0.87,0.92)** | **(2.10,2.21)** |
|  | 39.28 | 0.36 | 0.91 | 40.85 | 1.1 | 2.7 | 42.37 | 1.28 | 3.01 |
| Acre (NO) |  | (0.08,1.06) | (0.20,2.70) |  | (0.64,1.78) | (1.56,4.35) |  | (0.76,2.03) | (1.79,4.80) |
|  | 37.37 | 0.16 | 0.44 | 40.36 | 0.5 | 1.23 | 40.14 | 0.38 | 0.94 |
| Alagoas (NE) |  | (0.08,0.30) | (0.21,0.80) |  | (0.36,0.67) | (0.88,1.66) |  | (0.27,0.51) | (0.67,1.27) |
|  | 37.59 | 1.34 | 3.56 | 40.94 | 0.97 | 2.37 | 40.95 | 0.96 | 2.34 |
| Amapá (NO) |  | (0.47,3.08) | (1.24,8.20) |  | (0.51,1.67) | (1.25,4.08) |  | (0.53,1.58) | (1.30,3.86) |
|  | 37.94 | 1.68 | 4.42 | 41.67 | 0.85 | 2.04 | 41.54 | 0.62 | 1.49 |
| Amazonas (NO) |  | (1.23,2.24) | (3.23,5.91) |  | (0.65,1.10) | (1.55,2.64) |  | (0.46,0.81) | (1.10,1.95) |
|  | 38.57 | 0.37 | 0.95 | 41.38 | 0.48 | 1.16 | 41.47 | 0.46 | 1.11 |
| Bahia (NE) |  | (0.29,0.45) | (0.76,1.17) |  | (0.42,0.55) | (1.01,1.34) |  | (0.40,0.53) | (0.97,1.27) |
|  | 39.27 | 0.61 | 1.56 | 41.94 | 0.96 | 2.3 | 42.26 | 0.81 | 1.91 |
| Ceará (NE) |  | (0.49,0.76) | (1.25,1.94) |  | (0.84,1.11) | (1.99,2.64) |  | (0.70,0.93) | (1.65,2.20) |
|  | 39.75 | 1.37 | 3.44 | 43.37 | 1.11 | 2.56 | 43.64 | 0.89 | 2.04 |
| Distrito Federal (CW) |  | (0.94,1.92) | (2.36,4.83) |  | (0.84,1.44) | (1.94,3.33) |  | (0.67,1.15) | (1.54,2.64) |
|  | 38.34 | 1.08 | 2.81 | 42.04 | 1.15 | 2.74 | 42.23 | 0.84 | 1.99 |
| E. Santo (SE) |  | (0.85,1.35) | (2.21,3.53) |  | (0.96,1.38) | (2.28,3.28) |  | (0.69,1.02) | (1.63,2.42) |
|  | 37.92 | 1.09 | 2.88 | 41.6 | 1.35 | 3.24 | 41.85 | 1.08 | 2.57 |
| Goiás (CW) |  | (0.88,1.34) | (2.32,3.54) |  | (1.17,1.54) | (2.82,3.69) |  | (0.94,1.23) | (2.24,2.95) |
|  | 38.66 | 0.32 | 0.82 | 41.76 | 0.44 | 1.06 | 41.43 | 0.42 | 1.01 |
| Maranhão (NE) |  | (0.21,0.46) | (0.55,1.19) |  | (0.35,0.55) | (0.83,1.32) |  | (0.33,0.53) | (0.80,1.27) |
|  | 38.98 | 0.99 | 2.55 | 41.3 | 0.91 | 2.21 | 40.74 | 0.98 | 2.4 |
| Mato G. do Sul (CW) |  | (0.71,1.36) | (1.81,3.50) |  | (0.71,1.15) | (1.71,2.80) |  | (0.78,1.21) | (1.91,2.98) |
| Mato Grosso (CW) | 37.95 | 1.02 | 2.69 | 41.36 | 0.93 | 2.26 | 41.23 | 0.95 | 2.3 |
|  |  | (0.73,1.40) | (1.92,3.69) |  | (0.74,1.16) | (1.79,2.81) |  | (0.76,1.16) | (1.85,2.82) |
|  | 38.06 | 0.94 | 2.47 | 41.95 | 0.91 | 2.18 | 42.25 | 0.72 | 1.71 |
| Minas Gerais (SE) |  | (0.85,1.05) | (2.22,2.75) |  | (0.84,0.99) | (2.01,2.37) |  | (0.66,0.78) | (1.56,1.86) |
|  | 38.49 | 0.6 | 1.55 | 41.9 | 0.56 | 1.33 | 42.27 | 0.53 | 1.26 |
| Pará (NO) |  | (0.44,0.79) | (1.15,2.04) |  | (0.45,0.68) | (1.08,1.62) |  | (0.44,0.64) | (1.03,1.52) |
|  | 39.56 | 0.23 | 0.59 | 41.19 | 0.66 | 1.61 | 41.75 | 0.74 | 1.76 |
| Paraíba (NE) |  | (0.14,0.36) | (0.35,0.92) |  | (0.52,0.83) | (1.26,2.02) |  | (0.59,0.91) | (1.41,2.17) |
|  | 38.06 | 1.72 | 4.51 | 41.44 | 1.38 | 3.33 | 41.44 | 1.07 | 2.58 |
| Paraná (SO) |  | (1.53,1.92) | (4.02,5.05) |  | (1.25,1.51) | (3.03,3.66) |  | (0.97,1.18) | (2.34,2.85) |
|  | 36.67 | 0.75 | 2.05 | 40.38 | 0.72 | 1.79 | 40.66 | 0.56 | 1.38 |
| Pernambuco (NE) |  | (0.63,0.89) | (1.71,2.42) |  | (0.63,0.83) | (1.55,2.05) |  | (0.48,0.66) | (1.18,1.62) |
|  | 39.14 | 0.48 | 1.23 | 42.29 | 0.52 | 1.23 | 42.04 | 0.43 | 1.02 |
| Piauí (NE) |  | (0.29,0.76) | (0.73,1.95) |  | (0.38,0.69) | (0.90,1.64) |  | (0.31,0.57) | (0.74,1.37) |
|  | 36.26 | 2.21 | 6.1 | 39.97 | 0.95 | 2.38 | 40.8 | 0.73 | 1.8 |
| R. de Janeiro (SE) |  | (2.05,2.38) | (5.66,6.57) |  | (0.88,1.03) | (2.19,2.58) |  | (0.67,0.80) | (1.64,1.96) |
|  | 38.96 | 0.25 | 0.63 | 41.52 | 0.95 | 2.29 | 42.58 | 0.72 | 1.7 |
| Rio G. do Norte (NE) |  | (0.13,0.43) | (0.33,1.10) |  | (0.75,1.18) | (1.81,2.84) |  | (0.57,0.91) | (1.33,2.14) |
|  | 37.54 | 3.65 | 9.72 | 41.08 | 2.34 | 5.69 | 41.51 | 1.9 | 4.59 |
| Rio G. do Sul (SO) |  | (3.38,3.94) | (8.99,10.49) |  | (2.17,2.51) | (5.29,6.11) |  | (1.76,2.05) | (4.25,4.94) |
|  | 37.14 | 1.02 | 2.74 | 42.09 | 0.68 | 1.62 | 41.5 | 0.67 | 1.62 |
| Rondônia (NO) |  | (0.60,1.61) | (1.60,4.34) |  | (0.46,0.99) | (1.09,2.34) |  | (0.47,0.94) | (1.12,2.27) |
|  | 37.99 | 0.7 | 1.84 | 40.69 | 0.58 | 1.42 | 41.51 | 0.78 | 1.88 |
| Roraima (NO) |  | (0.14,2.13) | (0.37,5.62) |  | (0.22,1.25) | (0.54,3.08) |  | (0.39,1.41) | (0.94,3.40) |
|  | 38.24 | 2.45 | 6.4 | 42.29 | 1.79 | 4.24 | 42.58 | 1.85 | 4.34 |
| S. Catarina (SO) |  | (2.14,2.78) | (5.61,7.27) |  | (1.60,2.00) | (3.78,4.74) |  | (1.66,2.05) | (3.91,4.82) |
|  | 37.27 | 1.85 | 4.95 | 41.15 | 1.22 | 2.96 | 41.25 | 0.98 | 2.37 |
| S. Paulo (SE) |  | (1.75,1.95) | (4.69,5.22) |  | (1.16,1.28) | (2.81,3.10) |  | (0.93,1.03) | (2.25,2.50) |
|  | 38.05 | 0.83 | 2.18 | 41.14 | 0.6 | 1.47 | 41.58 | 0.72 | 1.73 |
| Sergipe (NE) |  | (0.55,1.20) | (1.44,3.16) |  | (0.42,0.84) | (1.03,2.04) |  | (0.52,0.98) | (1.25,2.35) |
|  | 39.4 | 0.38 | 0.97 | 42.66 | 0.74 | 1.74 | 42.21 | 0.91 | 2.16 |
| Tocantins (NO) |  | (0.14,0.85) | (0.35,2.15) |  | (0.49,1.08) | (1.14,2.53) |  | (0.62,1.29) | (1.47,3.04) |

**Note:** States are ordered alphabetically. 95% confidence intervals in parentheses. NO = North, NE = Northeast, CW = Central-West, SE = Southeast, SO = South. **Source:** SIM/DATASUS/MS and IBGE.

**Tables D2.**
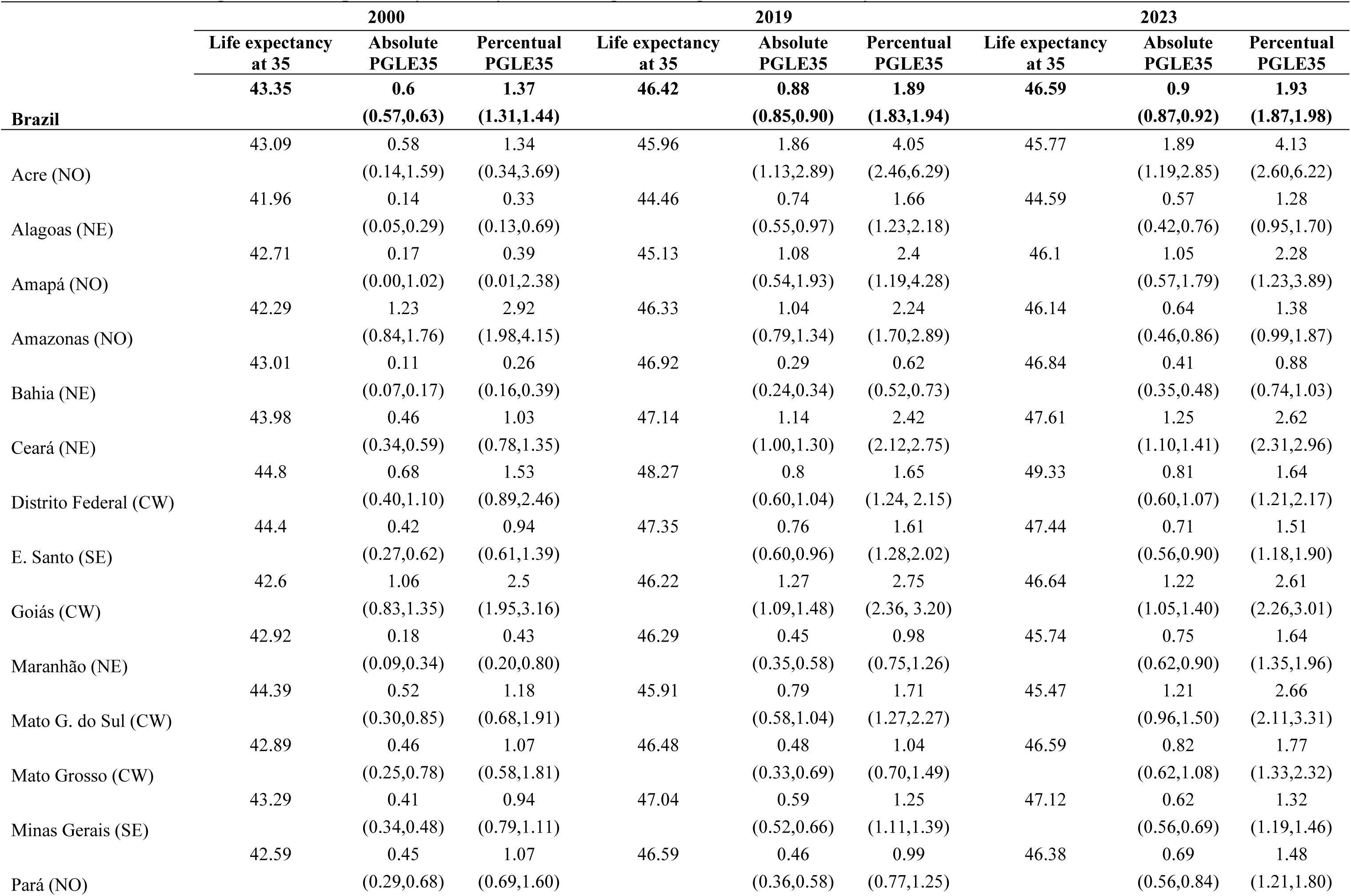
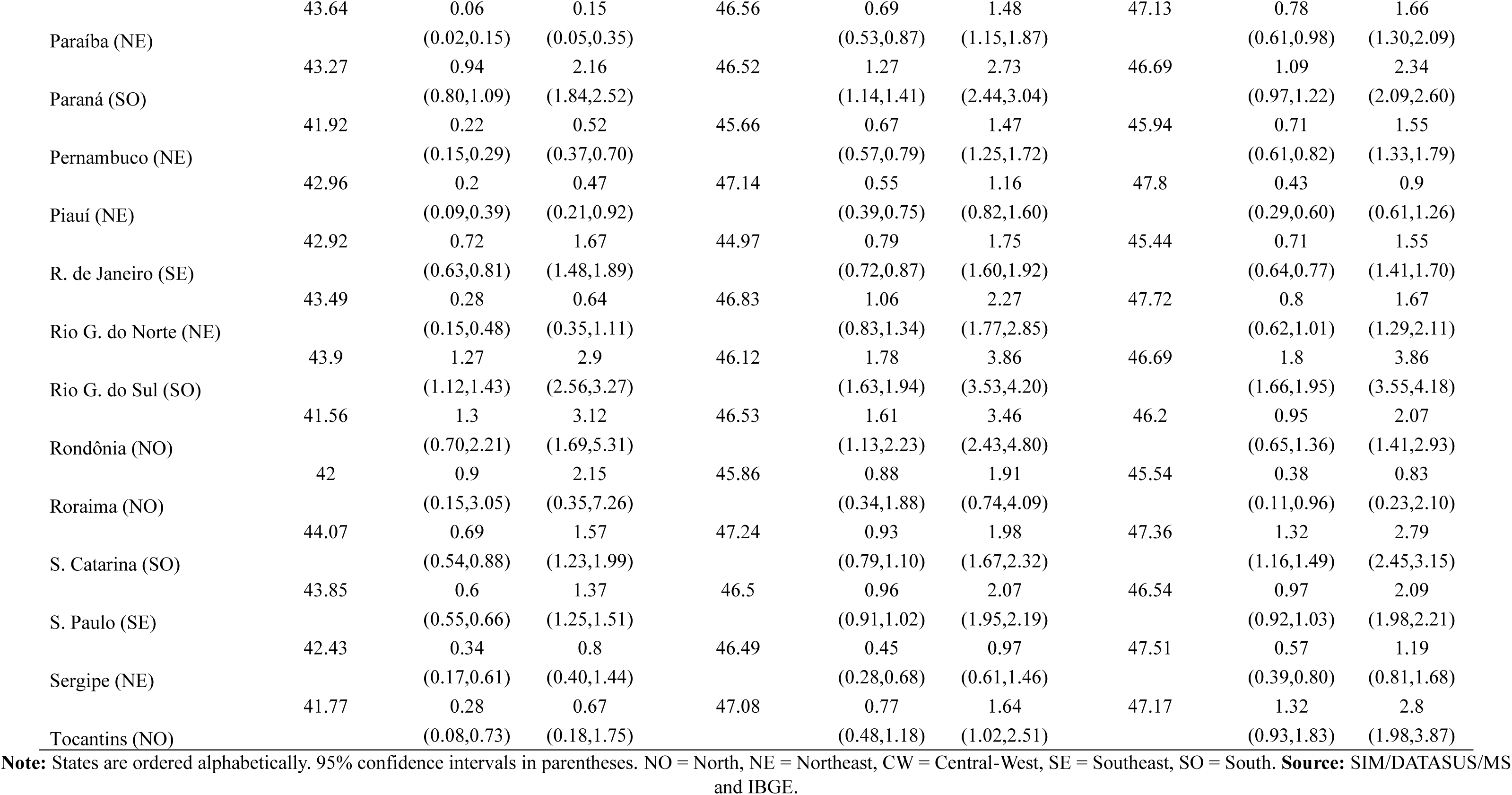
Potential gain in life expectancy at 35 by eliminating smoking-related mortality – Women, Brazilian states (2000, 2019 and 2023)

|  | 2000 |  |  | 2019 |  |  | 2023 |  |  |
| --- | --- | --- | --- | --- | --- | --- | --- | --- | --- |
|  | Life expectancy<br>at 35 | Absolute<br>PGLE35 | Percentual<br>PGLE35 | Life expectancy<br>at 35 | Absolute<br>PGLE35 | Percentual<br>PGLE35 | Life expectancy<br>at 35 | Absolute<br>PGLE35 | Percentual<br>PGLE35 |
|  | 43.35 | 0.6 | 1.37 | 46.42 | 0.88 | 1.89 | 46.59 | 0.9 | 1.93 |
| **Brazil** |  | **(0.57,0.63)** | **(1.31,1.44)** |  | **(0.85,0.90)** | **(1.83,1.94)** |  | **(0.87,0.92)** | **(1.87,1.98)** |
|  | 43.09 | 0.58 | 1.34 | 45.96 | 1.86 | 4.05 | 45.77 | 1.89 | 4.13 |
| Acre (NO) |  | (0.14,1.59) | (0.34,3.69) |  | (1.13,2.89) | (2.46,6.29) |  | (1.19,2.85) | (2.60,6.22) |
|  | 41.96 | 0.14 | 0.33 | 44.46 | 0.74 | 1.66 | 44.59 | 0.57 | 1.28 |
| Alagoas (NE) |  | (0.05,0.29) | (0.13,0.69) |  | (0.55,0.97) | (1.23,2.18) |  | (0.42,0.76) | (0.95,1.70) |
|  | 42.71 | 0.17 | 0.39 | 45.13 | 1.08 | 2.4 | 46.1 | 1.05 | 2.28 |
| Amapá (NO) |  | (0.00,1.02) | (0.01,2.38) |  | (0.54,1.93) | (1.19,4.28) |  | (0.57,1.79) | (1.23,3.89) |
|  | 42.29 | 1.23 | 2.92 | 46.33 | 1.04 | 2.24 | 46.14 | 0.64 | 1.38 |
| Amazonas (NO) |  | (0.84,1.76) | (1.98,4.15) |  | (0.79,1.34) | (1.70,2.89) |  | (0.46,0.86) | (0.99,1.87) |
|  | 43.01 | 0.11 | 0.26 | 46.92 | 0.29 | 0.62 | 46.84 | 0.41 | 0.88 |
| Bahia (NE) |  | (0.07,0.17) | (0.16,0.39) |  | (0.24,0.34) | (0.52,0.73) |  | (0.35,0.48) | (0.74,1.03) |
|  | 43.98 | 0.46 | 1.03 | 47.14 | 1.14 | 2.42 | 47.61 | 1.25 | 2.62 |
| Ceará (NE) |  | (0.34,0.59) | (0.78,1.35) |  | (1.00,1.30) | (2.12,2.75) |  | (1.10,1.41) | (2.31,2.96) |
|  | 44.8 | 0.68 | 1.53 | 48.27 | 0.8 | 1.65 | 49.33 | 0.81 | 1.64 |
| Distrito Federal (CW) |  | (0.40,1.10) | (0.89,2.46) |  | (0.60,1.04) | (1.24, 2.15) |  | (0.60,1.07) | (1.21,2.17) |
|  | 44.4 | 0.42 | 0.94 | 47.35 | 0.76 | 1.61 | 47.44 | 0.71 | 1.51 |
| E. Santo (SE) |  | (0.27,0.62) | (0.61,1.39) |  | (0.60,0.96) | (1.28,2.02) |  | (0.56,0.90) | (1.18,1.90) |
|  | 42.6 | 1.06 | 2.5 | 46.22 | 1.27 | 2.75 | 46.64 | 1.22 | 2.61 |
| Goiás (CW) |  | (0.83,1.35) | (1.95,3.16) |  | (1.09,1.48) | (2.36, 3.20) |  | (1.05,1.40) | (2.26,3.01) |
|  | 42.92 | 0.18 | 0.43 | 46.29 | 0.45 | 0.98 | 45.74 | 0.75 | 1.64 |
| Maranhão (NE) |  | (0.09,0.34) | (0.20,0.80) |  | (0.35,0.58) | (0.75,1.26) |  | (0.62,0.90) | (1.35,1.96) |
|  | 44.39 | 0.52 | 1.18 | 45.91 | 0.79 | 1.71 | 45.47 | 1.21 | 2.66 |
| Mato G. do Sul (CW) |  | (0.30,0.85) | (0.68,1.91) |  | (0.58,1.04) | (1.27,2.27) |  | (0.96,1.50) | (2.11,3.31) |
|  | 42.89 | 0.46 | 1.07 | 46.48 | 0.48 | 1.04 | 46.59 | 0.82 | 1.77 |
| Mato Grosso (CW) |  | (0.25,0.78) | (0.58,1.81) |  | (0.33,0.69) | (0.70,1.49) |  | (0.62,1.08) | (1.33,2.32) |
|  | 43.29 | 0.41 | 0.94 | 47.04 | 0.59 | 1.25 | 47.12 | 0.62 | 1.32 |
| Minas Gerais (SE) |  | (0.34,0.48) | (0.79,1.11) |  | (0.52,0.66) | (1.11,1.39) |  | (0.56,0.69) | (1.19,1.46) |
|  | 42.59 | 0.45 | 1.07 | 46.59 | 0.46 | 0.99 | 46.38 | 0.69 | 1.48 |
| Pará (NO) |  | (0.29,0.68) | (0.69,1.60) |  | (0.36,0.58) | (0.77,1.25) |  | (0.56,0.84) | (1.21,1.80) |
|  | 43.64 | 0.06 | 0.15 | 46.56 | 0.69 | 1.48 | 47.13 | 0.78 | 1.66 |
| Paraíba (NE) |  | (0.02,0.15) | (0.05,0.35) |  | (0.53,0.87) | (1.15,1.87) |  | (0.61,0.98) | (1.30,2.09) |
|  | 43.27 | 0.94 | 2.16 | 46.52 | 1.27 | 2.73 | 46.69 | 1.09 | 2.34 |
| Paraná (SO) |  | (0.80,1.09) | (1.84,2.52) |  | (1.14,1.41) | (2.44,3.04) |  | (0.97,1.22) | (2.09,2.60) |
|  | 41.92 | 0.22 | 0.52 | 45.66 | 0.67 | 1.47 | 45.94 | 0.71 | 1.55 |
| Pernambuco (NE) |  | (0.15,0.29) | (0.37,0.70) |  | (0.57,0.79) | (1.25,1.72) |  | (0.61,0.82) | (1.33,1.79) |
|  | 42.96 | 0.2 | 0.47 | 47.14 | 0.55 | 1.16 | 47.8 | 0.43 | 0.9 |
| Piauí (NE) |  | (0.09,0.39) | (0.21,0.92) |  | (0.39,0.75) | (0.82,1.60) |  | (0.29,0.60) | (0.61,1.26) |
|  | 42.92 | 0.72 | 1.67 | 44.97 | 0.79 | 1.75 | 45.44 | 0.71 | 1.55 |
| R. de Janeiro (SE) |  | (0.63,0.81) | (1.48,1.89) |  | (0.72,0.87) | (1.60,1.92) |  | (0.64,0.77) | (1.41,1.70) |
|  | 43.49 | 0.28 | 0.64 | 46.83 | 1.06 | 2.27 | 47.72 | 0.8 | 1.67 |
| Rio G. do Norte (NE) |  | (0.15,0.48) | (0.35,1.11) |  | (0.83,1.34) | (1.77,2.85) |  | (0.62,1.01) | (1.29,2.11) |
|  | 43.9 | 1.27 | 2.9 | 46.12 | 1.78 | 3.86 | 46.69 | 1.8 | 3.86 |
| Rio G. do Sul (SO) |  | (1.12,1.43) | (2.56,3.27) |  | (1.63,1.94) | (3.53,4.20) |  | (1.66,1.95) | (3.55,4.18) |
|  | 41.56 | 1.3 | 3.12 | 46.53 | 1.61 | 3.46 | 46.2 | 0.95 | 2.07 |
| Rondônia (NO) |  | (0.70,2.21) | (1.69,5.31) |  | (1.13,2.23) | (2.43,4.80) |  | (0.65,1.36) | (1.41,2.93) |
|  | 42 | 0.9 | 2.15 | 45.86 | 0.88 | 1.91 | 45.54 | 0.38 | 0.83 |
| Roraima (NO) |  | (0.15,3.05) | (0.35,7.26) |  | (0.34,1.88) | (0.74,4.09) |  | (0.11,0.96) | (0.23,2.10) |
|  | 44.07 | 0.69 | 1.57 | 47.24 | 0.93 | 1.98 | 47.36 | 1.32 | 2.79 |
| S. Catarina (SO) |  | (0.54,0.88) | (1.23,1.99) |  | (0.79,1.10) | (1.67,2.32) |  | (1.16,1.49) | (2.45,3.15) |
|  | 43.85 | 0.6 | 1.37 | 46.5 | 0.96 | 2.07 | 46.54 | 0.97 | 2.09 |
| S. Paulo (SE) |  | (0.55,0.66) | (1.25,1.51) |  | (0.91,1.02) | (1.95,2.19) |  | (0.92,1.03) | (1.98,2.21) |
|  | 42.43 | 0.34 | 0.8 | 46.49 | 0.45 | 0.97 | 47.51 | 0.57 | 1.19 |
| Sergipe (NE) |  | (0.17,0.61) | (0.40,1.44) |  | (0.28,0.68) | (0.61,1.46) |  | (0.39,0.80) | (0.81,1.68) |
|  | 41.77 | 0.28 | 0.67 | 47.08 | 0.77 | 1.64 | 47.17 | 1.32 | 2.8 |
| Tocantins (NO) |  | (0.08,0.73) | (0.18,1.75) |  | (0.48,1.18) | (1.02,2.51) |  | (0.93,1.83) | (1.98,3.87) |

**Note:** States are ordered alphabetically. 95% confidence intervals in parentheses. NO = North, NE = Northeast, CW = Central-West, SE = Southeast, SO = South. **Source:** SIM/DATASUS/MS and IBGE.

**Table D3.** Age- standardized smoking-related mortality rates - Men, Brazilian states (2000, 2019 and

|  | **2000** | **2019** | **2023** |
| --- | --- | --- | --- |
|  | **19.37** | **11.47** | **9.44** |
| **Brazil** | **(18.82,19.93)** | **(11.20,11.74)** | **(9.21,9.67)** |
|  | 4.29 | 12.27 | 12.13 |
| Acre (NO) | (0.97,12.45) | (7.36,19.35) | (7.39,18.94) |
|  | 1.91 | 5.63 | 4.54 |
| Alagoas (NE) | (0.89,3.64) | (4.07,7.63) | (3.29,6.11) |
|  | 18.41 | 10.68 | 10.81 |
| Amapá (NO) | (7.47,38.24) | (5.77,18.06) | (6.26,17.44) |
|  | 20.97 | 9.12 | 6.62 |
| Amazonas (NO) | (15.66,27.52) | (7.04,11.61) | (4.96,8.64) |
|  | 4.21 | 5.08 | 4.99 |
| Bahia (NE) | (3.37,5.19) | (4.41,5.83) | (4.36,5.67) |
|  | 6.85 | 9.41 | 7.55 |
| Ceará (NE) | (5.48,8.45) | (8.17,10.76) | (6.56,8.66) |
|  | 15.43 | 10.32 | 8.37 |
| Distrito Federal (CW) | (10.86,21.14) | (7.98,13.12) | (6.44,10.62) |
|  | 13 | 11.53 | 8.32 |
| E. Santo (SE) | (10.20,16.30) | (9.66,13.64) | (6.85,10.00) |
|  | 13.94 | 13.69 | 10.72 |
| Goiás (CW) | (11.36,16.92) | (12.09,15.47) | (9.41,12.15) |
|  | 3.56 | 5.01 | 5.17 |
| Maranhão (NE) | (2.35,5.18) | (3.99,6.19) | (4.17,6.37) |
|  | 11.56 | 9.71 | 11.03 |
| Mato G. do Sul (CW) | (8.31,15.70) | (7.59,12.17) | (8.91,13.54) |
|  | 12.86 | 9.97 | 10.29 |
| Mato Grosso (CW) | (9.31,17.44) | (7.96,12.30) | (8.40,12.51) |
|  | 12.01 | 9.12 | 6.99 |
| Minas Gerais (SE) | (10.77,13.33) | (8.43,9.85) | (6.42,7.60) |
|  | 7.33 | 5.77 | 5.6 |
| Pará (NO) | (5.42,9.72) | (4.71,7.03) | (4.63,6.71) |
|  | 2.43 | 7.02 | 7.17 |
| Paraíba (NE) | (1.43,3.81) | (5.56,8.77) | (5.74,8.81) |
|  | 22.42 | 14.93 | 11.42 |
| Paraná (SO) | (20.13,24.92) | (13.67,16.26) | (10.38,12.53) |
|  | 9.62 | 8.1 | 6.08 |
| Pernambuco (NE) | (8.04,11.44) | (7.06,9.25) | (5.19,7.10) |
|  | 5.55 | 5.16 | 4.16 |
| Piauí (NE) | (3.29,8.75) | (3.81,6.84) | (3.04,5.58) |
|  | 29.89 | 11.34 | 8.24 |
| R. de Janeiro (SE) | (27.85,32.09) | (10.46,12.26) | (7.54,8.99) |
|  | 2.92 | 9.67 | 6.87 |
| Rio G. do Norte (NE) | (1.51,5.10) | (7.76,11.91) | (5.42,8.57) |
|  | 46.34 | 24.71 | 19.74 |
| Rio G. do Sul (SO) | (43.24,49.57) | (23.18,26.30) | (18.46,21.10) |
|  | 13.41 | 7.12 | 7.13 |
| Rondônia (NO) | (7.95,21.02) | (4.83,10.04) | (4.91,9.90) |
|  | 8.2 | 6.7 | 8.31 |
| Roraima (NO) | (1.77,24.13) | (2.60,14.17) | (4.26,14.72) |
| S. Catarina (SO) | 31.3 | 18.01 | 17.74 |
|  | (27.68,35.30) | (16.21,19.89) | (16.13,19.53) |
|  | 24.59 | 13.35 | 10.95 |
| S. Paulo (SE) | (23.37,25.87) | (12.75,13.98) | (10.44,11.48) |
|  | 9.62 | 6.56 | 7.39 |
| Sergipe (NE) | (6.29,14.02) | (4.61,9.04) | (5.39,9.94) |
|  | 4.49 | 6.83 | 8.59 |
| Tocantins (NO) | (1.67,9.89) | (4.59,9.85) | (5.97,12.01) |

**Note:** States are ordered alphabetically. 95% confidence intervals in parentheses. NO = North, NE = Northeast, CW = Central-West, SE = Southeast, SO = South. **Source:** SIM/DATASUS/MS and IBGE.

**Table D4.** Age- standardized smoking-related mortality rates – Women, Brazilian states (2000, 2019 and 2023)

|  | 2000 | 2019 | 2023 |
| --- | --- | --- | --- |
|  | 5.55 | 6.6 | 6.64 |
| **Brazil** | **(5.28,5.83)** | **(6.41,6.80)** | **(6.46,6.82)** |
|  | 5.42 | 13.03 | 13.92 |
| Acre (NO) | (1.42,14.63) | (8.12,19.91) | (9.13,20.48) |
|  | 1.33 | 6.58 | 5.12 |
| Alagoas (NE) | (0.50,2.83) | (4.92,8.60) | (3.79,6.77) |
|  | 1.48 | 9.04 | 8.4 |
| Amapá (NO) | (0.03,8.65) | (4.62,16.17) | (4.80,13.67) |
|  | 11.87 | 7.31 | 5.11 |
| Amazonas (NO) | (8.14,16.70) | (5.57,9.43) | (3.70,6.83) |
|  | 1.02 | 2.24 | 3.13 |
| Bahia (NE) | (0.64,1.56) | (1.88,2.65) | (2.67,3.66) |
|  | 4.09 | 7.94 | 7.99 |
| Ceará (NE) | (3.09,5.36) | (6.97,9.00) | (7.03,9.04) |
|  | 5.72 | 5.41 | 4.67 |
| Distrito Federal (CW) | (3.44,8.93) | (4.12,7.00) | (3.44,6.24) |
|  | 3.37 | 4.98 | 4.68 |
| E. Santo (SE) | (2.19,4.94) | (3.91,6.25) | (3.60,5.97) |
|  | 10.49 | 9.18 | 8.7 |
| Goiás (CW) | (8.31,13.21) | (7.92,10.56) | (7.57,9.98) |
|  | 1.66 | 3.59 | 7.03 |
| Maranhão (NE) | (0.79,3.12) | (2.74,4.64) | (5.92,8.29) |
|  | 4.36 | 6.35 | 9.32 |
| Mato G. do Sul (CW) | (2.57,7.02) | (4.73,8.39) | (7.51,11.50) |
|  | 4.69 | 3.71 | 6.09 |
| Mato Grosso (CW) | (2.62,7.74) | (2.51,5.26) | (4.58,7.92) |
|  | 3.78 | 4.06 | 4.45 |
| Minas Gerais (SE) | (3.17,4.43) | (3.62,4.55) | (4.02,4.93) |
|  | 4.44 | 3.42 | 5.32 |
| Pará (NO) | (2.90,6.55) | (2.68,4.31) | (4.34,6.43) |
|  | 0.57 | 5 | 5.58 |
| Paraíba (NE) | (0.17,1.44) | (3.88,6.35) | (4.39,7.00) |
|  | 8.89 | 9.53 | 8 |
| Paraná (SO) | (7.60,10.35) | (8.55,10.57) | (7.15,8.90) |
|  | 2.13 | 5.29 | 5.35 |
| Pernambuco (NE) | (1.52,2.90) | (4.48,6.20) | (4.60,6.19) |
|  | 1.98 | 4.17 | 3.01 |
| Piauí (NE) | (0.90,3.84) | (2.97,5.70) | (2.05,4.24) |
|  | 6.78 | 6.6 | 5.73 |
| R. de Janeiro (SE) | (5.99,7.65) | (6.00,7.23) | (5.21,6.27) |
|  | 2.54 | 7.73 | 5.56 |
| Rio G. do Norte (NE) | (1.35,4.33) | (6.04,9.69) | (4.33,7.01) |
|  | 11.49 | 13.58 | 12.79 |
| Rio G. do Sul (SO) | (10.18,12.92) | (12.49,14.74) | (11.79,13.85) |
|  | 13.48 | 10.07 | 6.87 |
| Rondônia (NO) | (7.49,22.22) | (7.17,13.70) | (4.71,9.72) |
|  | 8.65 | 7.09 | 3.03 |
| Roraima (NO) | (1.60,27.36) | (2.85,14.76) | (0.86,7.90) |
|  | 6.34 | 6.95 | 8.96 |
| S. Catarina (SO) | (4.98,7.96) | (5.91,8.15) | (7.92,10.09) |
|  | 5.53 | 7.28 | 7.32 |
| S. Paulo (SE) | (5.03,6.07) | (6.86,7.71) | (6.93,7.72) |
|  | 3.24 | 3.5 | 3.64 |
| Sergipe (NE) | (1.61,5.83) | (2.20,5.29) | (2.46,5.19) |
|  | 2.73 | 5.03 | 7.98 |
| Tocantins (NO) | (0.69,7.34) | (3.08,7.78) | (5.60,10.96) |

**Note:** States are ordered alphabetically. 95% confidence intervals in parentheses. NO = North, NE = Northeast, CW = Central-West, SE = Southeast, SO = South. **Source:** SIM/DATASUS/MS and IBGE.

**Table D5.** Total smoking-related contribution to the change in life expectancy at age 35– Men, Brazilian states (2000 to 2019 and 2000 to 2023)

|  | 2000-2019 |  |  | 2000-2023 |  |  |
| --- | --- | --- | --- | --- | --- | --- |
|  | Change in life expectancy at 35 | Absolute contribution | Relative contribution (%) | Change in life expectancy at 35 | Absolute contribution | Percentual contribution (%) |
|  | **3.51** | **0.71** | **20.13** | **3.74** | **0.88** | **23.5** |
| **Brazil** |  | **(0.68, 0.73)** | **(19.47, 20.80)** |  | **(0.85, 0.90)** | **(22.78, 24.22)** |
|  | 1.57 | -0.65 | -41.2 | 3.08 | -0.7 | -22.69 |
| Acre (NO) |  | (-0.66, -0.63) | (-42.19, -40.22) |  | (-0.73, -0.67) | (-23.72, -21.66) |
|  | 2.99 | -0.27 | -9.01 | 2.77 | -0.16 | -5.75 |
| Alagoas (NE) |  | (-0.29, -0.25) | (-9.66, -8.38) |  | (-0.17, -0.15) | (-6.20, -5.31) |
|  | 3.35 | 0.49 | 14.5 | 3.35 | 0.51 | 15.14 |
| Amapá (NO) |  | (-0.21, 1.19) | (-6.30, 35.62) |  | (-0.23, 1.26) | (-6.80, 37.47) |
|  | 3.73 | 1.05 | 28.07 | 3.6 | 1.23 | 34.32 |
| Amazonas (NO) |  | (0.73, 1.36) | (19.57, 36.50) |  | (0.87, 1.59) | (24.33, 44.26) |
|  | 2.81 | -0.05 | -1.64 | 2.9 | -0.03 | -0.87 |
| Bahia (NE) |  | (-0.07, -0.03) | (-2.36, -0.92) |  | (-0.05, 0.00) | (-1.76, 0.04) |
|  | 2.68 | -0.19 | -7.15 | 2.99 | -0.03 | -1.02 |
| Ceará (NE) |  | (-0.22, -0.17) | (-8.05, -6.24) |  | (-0.08, 0.02) | (-2.54, 0.52) |
|  | 3.61 | 0.47 | 13.11 | 3.89 | 0.67 | 17.15 |
| Distrito Federal (CW) |  | (0.24, 0.71) | (6.69, 19.61) |  | (0.39, 0.95) | (9.91, 24.44) |
|  | 3.71 | 0.22 | 5.82 | 3.89 | 0.47 | 12.1 |
| E. Santo (SE) |  | (0.11, 0.32) | (3.07, 8.57) |  | (0.34, 0.61) | (8.61, 15.56) |
|  | 3.68 | 0.08 | 2.28 | 3.93 | 0.32 | 8.17 |
| Goiás (CW) |  | (-0.01, 0.18) | (-0.32, 4.83) |  | (0.20, 0.44) | (5.11, 11.23) |
|  | 3.1 | -0.06 | -1.96 | 2.77 | -0.06 | -2.32 |
| Maranhão (NE) |  | (-0.10, -0.02) | (-3.28, -0.65) |  | (-0.11, -0.02) | (-3.80, -0.86) |
|  | 2.31 | 0.19 | 8.17 | 1.76 | 0.1 | 5.41 |
| Mato G. do Sul (CW) |  | (0.07, 0.31) | (2.93, 13.30) |  | (-0.02, 0.21) | (-1.35, 12.21) |
|  | 3.41 | 0.28 | 8.24 | 3.28 | 0.26 | 7.79 |
| Mato Grosso (CW) |  | (0.13, 0.44) | (3.68, 12.88) |  | (0.09, 0.42) | (2.74, 12.94) |
|  | 3.89 | 0.28 | 7.32 | 4.19 | 0.45 | 10.84 |
| Minas Gerais (SE) |  | (0.24, 0.33) | (6.06, 8.57) |  | (0.39, 0.52) | (9.39, 12.30) |
|  | 3.41 | 0.16 | 4.7 | 3.77 | 0.2 | 5.36 |
| Pará (NO) |  | (0.07, 0.25) | (2.17, 7.21) |  | (0.10, 0.30) | (2.74, 7.97) |
| Paraíba (NE) | 1.63 | -0.37 | -22.51 | 2.19 | -0.39 | -17.94 |
|  |  | (-0.39, -0.34) | (-24.04, -20.98) |  | (-0.41, -0.37) | (-18.84, -17.05) |
|  | 3.38 | 0.67 | 19.74 | 3.38 | 0.92 | 27.31 |
| Paraná (SO) |  | (0.58, 0.76) | (17.07, 22.40) |  | (0.82, 1.03) | (24.12, 30.50) |
|  | 3.71 | 0.19 | 5.23 | 3.99 | 0.35 | 8.68 |
| Pernambuco (NE) |  | (0.14, 0.25) | (3.79, 6.66) |  | (0.28, 0.41) | (7.02, 10.33) |
|  | 3.15 | 0.07 | 2.35 | 2.89 | 0.15 | 5.1 |
| Piauí (NE) |  | (-0.05, 0.19) | (-1.52, 6.11) |  | (0.01, 0.29) | (0.25, 9.96) |
|  | 3.72 | 1.47 | 39.62 | 4.55 | 1.75 | 38.45 |
| R. de Janeiro (SE) |  | (1.38, 1.57) | (37.13, 42.12) |  | (1.64, 1.86) | (36.09, 40.84) |
|  | 2.56 | -0.55 | -21.58 | 3.63 | -0.35 | -9.66 |
| Rio G. do Norte (NE) |  | (-0.57, -0.54) | (-22.18, -20.98) |  | (-0.37, -0.33) | (-10.32, -9.00) |
|  | 3.54 | 1.8 | 50.78 | 3.96 | 2.21 | 55.74 |
| Rio G. do Sul (SO) |  | (1.67, 1.92) | (47.23, 54.34) |  | (2.06, 2.36) | (52.00, 59.46) |
|  | 4.95 | 0.58 | 11.66 | 4.36 | 0.54 | 12.37 |
| Rondônia (NO) |  | (0.23, 0.92) | (4.61, 18.61) |  | (0.19, 0.88) | (4.44, 20.21) |
|  | 2.71 | 0.2 | 7.35 | 3.52 | 0.06 | 1.6 |
| Roraima (NO) |  | (-0.33, 0.73) | (-12.31, 26.96) |  | (-0.55, 0.65) | (-15.49, 18.55) |
|  | 4.05 | 1.17 | 28.91 | 4.34 | 1.22 | 28.03 |
| S. Catarina (SO) |  | (1.01, 1.33) | (24.98, 32.88) |  | (1.05, 1.39) | (24.11, 32.08) |
|  | 3.87 | 0.93 | 23.88 | 3.98 | 1.12 | 28.15 |
| S. Paulo (SE) |  | (0.87, 0.98) | (22.54, 25.21) |  | (1.06, 1.18) | (26.68, 29.61) |
|  | 3.09 | 0.34 | 11.12 | 3.53 | 0.28 | 7.86 |
| Sergipe (NE) |  | (0.18, 0.50) | (5.79, 16.29) |  | (0.12, 0.44) | (3.35, 12.41) |
|  | 3.26 | -0.21 | -6.56 | 2.81 | -0.34 | -11.98 |
| Tocantins (NO) |  | (-0.35, -0.08) | (-10.71, -2.33) |  | (-0.45, -0.22) | (-16.00, -8.01) |

**Note:** States are ordered alphabetically. 95% confidence intervals in parentheses. NO = North, NE = Northeast, CW = Central-West, SE = Southeast, SO = South. **Source:** SIM/DATASUS/MS and IBGE.

**Table D6.** Total smoking-related contribution to the change in life expectancy at age 35– Women, Brazilian states (2000 to 2019 and 2000 to 2023)

|  | 2019-2000 |  |  | 2023-2000 |  |  |
| --- | --- | --- | --- | --- | --- | --- |
|  | Change in life expectancy at 35 | Absolute contribution | Percentual contribution (%) | Change in life expectancy at 35 | Absolute contribution | Percentual contribution (%) |
|  | **3.07** | **-0.12** | **-3.97** | **3.24** | **-0.13** | **-3.87** |
| **Brazil** |  | **(-0.13, -0.11)** | **(-4.27, -3.68)** |  | **(-0.14, -0.11)** | **(-4.20, -3.54)** |
|  | 2.88 | -0.92 | -31.93 | 2.68 | -0.96 | -35.71 |
| Acre (NO) |  | (-1.01, -0.83) | (-35.08, -28.70) |  | (-1.09, -0.83) | (-40.62, -30.84) |
|  | 2.5 | -0.54 | -21.48 | 2.63 | -0.38 | -14.47 |
| Alagoas (NE) |  | (-0.60, -0.47) | (-24.01, -18.97) |  | (-0.41, -0.35) | (-15.44, -13.49) |
|  | 2.42 | -0.8 | -33.15 | 3.39 | -0.7 | -20.55 |
| Amapá (NO) |  | (-1.06, -0.54) | (-43.69, -22.47) |  | (-1.11, -0.28) | (-32.86, -8.28) |
|  | 4.04 | 0.47 | 11.74 | 3.85 | 0.76 | 19.76 |
| Amazonas (NO) |  | (0.22, 0.73) | (5.40, 18.16) |  | (0.46, 1.06) | (11.99, 27.60) |
|  | 3.91 | -0.15 | -3.71 | 3.83 | -0.24 | -6.39 |
| Bahia (NE) |  | (-0.15, -0.14) | (-3.92, -3.50) |  | (-0.25, -0.24) | (-6.52, -6.26) |
|  | 3.17 | -0.48 | -15.14 | 3.64 | -0.5 | -13.87 |
| Ceará (NE) |  | (-0.49, -0.47) | (-15.47, -14.79) |  | (-0.52, -0.49) | (-14.17, -13.57) |
|  | 3.47 | 0.07 | 1.88 | 4.53 | 0.16 | 3.5 |
| Distrito Federal (CW) |  | (-0.11, 0.24) | (-3.15, 6.88) |  | (-0.03, 0.36) | (-0.71, 7.85) |
|  | 2.96 | -0.19 | -6.52 | 3.05 | -0.15 | -4.81 |
| E. Santo (SE) |  | (-0.23, -0.16) | (-7.62, -5.43) |  | (-0.18, -0.11) | (-5.95, -3.67) |
|  | 3.62 | 0.12 | 3.39 | 4.04 | 0.17 | 4.31 |
| Goiás (CW) |  | (0.00, 0.24) | (0.09, 6.66) |  | (0.04, 0.31) | (0.99, 7.59) |
|  | 3.36 | -0.23 | -6.82 | 2.81 | -0.56 | -20.07 |
| Maranhão (NE) |  | (-0.26, -0.20) | (-7.64, -6.02) |  | (-0.57, -0.56) | (-20.23, -19.92) |
|  | 1.51 | -0.24 | -16.03 | 1.08 | -0.61 | -55.93 |
| Mato G. do Sul (CW) |  | (-0.29, -0.20) | (-18.94, -13.06) |  | (-0.63, -0.58) | (-58.19, -53.68) |
|  | 3.6 | 0.08 | 2.19 | 3.7 | -0.19 | -5.23 |
| Mato Grosso (CW) |  | (-0.05, 0.20) | (-1.28, 5.62) |  | (-0.29, -0.10) | (-7.76, -2.67) |
|  | 3.75 | -0.03 | -0.93 | 3.83 | -0.07 | -1.85 |
| Minas Gerais (SE) |  | (-0.06, -0.01) | (-1.48, -0.37) |  | (-0.09, -0.05) | (-2.45, -1.24) |
|  | 4.01 | 0.1 | 2.57 | 3.79 | -0.1 | -2.71 |
| Pará (NO) |  | (-0.01, 0.21) | (-0.23, 5.33) |  | (-0.19, -0.02) | (-5.04, -0.41) |
| Paraíba (NE) | 2.91 | -0.53 | -18.34 | 3.48 | -0.6 | -17.18 |
|  |  | (-0.60, -0.46) | (-20.78, -15.91) |  | (-0.68, -0.52) | (-19.46, -14.85) |
|  | 3.25 | -0.06 | -1.93 | 3.43 | 0.1 | 3.01 |
| Paraná (SO) |  | (-0.10, -0.02) | (-3.19, -0.67) |  | (0.05, 0.16) | (1.33, 4.70) |
|  | 3.75 | -0.33 | -8.87 | 4.03 | -0.35 | -8.64 |
| Pernambuco (NE) |  | (-0.35, -0.32) | (-9.31, -8.43) |  | (-0.36, -0.34) | (-8.90, -8.38) |
|  | 4.18 | -0.25 | -6 | 4.84 | -0.13 | -2.69 |
| Piauí (NE) |  | (-0.26, -0.24) | (-6.25, -5.75) |  | (-0.17, -0.09) | (-3.54, -1.84) |
|  | 2.06 | 0.03 | 1.3 | 2.52 | 0.12 | 4.88 |
| R. de Janeiro (SE) |  | (0.00, 0.05) | (0.10, 2.51) |  | (0.09, 0.16) | (3.54, 6.25) |
|  | 3.34 | -0.61 | -18.22 | 4.23 | -0.36 | -8.57 |
| Rio G. do Norte (NE) |  | (-0.64, -0.57) | (-19.28, -17.15) |  | (-0.38, -0.34) | (-9.00, -8.14) |
|  | 2.22 | -0.22 | -9.87 | 2.79 | -0.14 | -5.18 |
| Rio G. do Sul (SO) |  | (-0.25, -0.19) | (-11.15, -8.58) |  | (-0.18, -0.11) | (-6.61, -3.77) |
|  | 4.97 | 0.33 | 6.58 | 4.64 | 0.7 | 15 |
| Rondônia (NO) |  | (-0.12, 0.77) | (-2.34, 15.50) |  | (0.17, 1.22) | (3.57, 26.20) |
|  | 3.86 | 0.19 | 4.86 | 3.53 | 0.61 | 17.21 |
| Roraima (NO) |  | (-0.70, 1.07) | (-18.15, 27.87) |  | (-0.54, 1.74) | (-15.17, 49.23) |
|  | 3.17 | -0.06 | -1.81 | 3.29 | -0.3 | -9.04 |
| S. Catarina (SO) |  | (-0.10, -0.01) | (-3.30, -0.31) |  | (-0.35, -0.25) | (-10.54, -7.50) |
|  | 2.65 | -0.2 | -7.58 | 2.69 | -0.2 | -7.54 |
| S. Paulo (SE) |  | (-0.21, -0.19) | (-8.01, -7.15) |  | (-0.22, -0.19) | (-8.10, -6.98) |
|  | 4.06 | -0.03 | -0.66 | 5.08 | -0.07 | -1.42 |
| Sergipe (NE) |  | (-0.10, 0.04) | (-2.35, 1.00) |  | (-0.16, 0.01) | (-3.05, 0.25) |
|  | 5.31 | -0.25 | -4.71 | 5.4 | -0.62 | -11.41 |
| Tocantins (NO) |  | (-0.38, -0.12) | (-7.15, -2.28) |  | (-0.70, -0.53) | (-12.94, -9.86) |

**Note:** States are ordered alphabetically. 95% confidence intervals in parentheses. NO = North, NE = Northeast, CW = Central-West, SE = Southeast, SO = South. **Source:** SIM/DATASUS/MS and IBGE.

**Figure D1.**
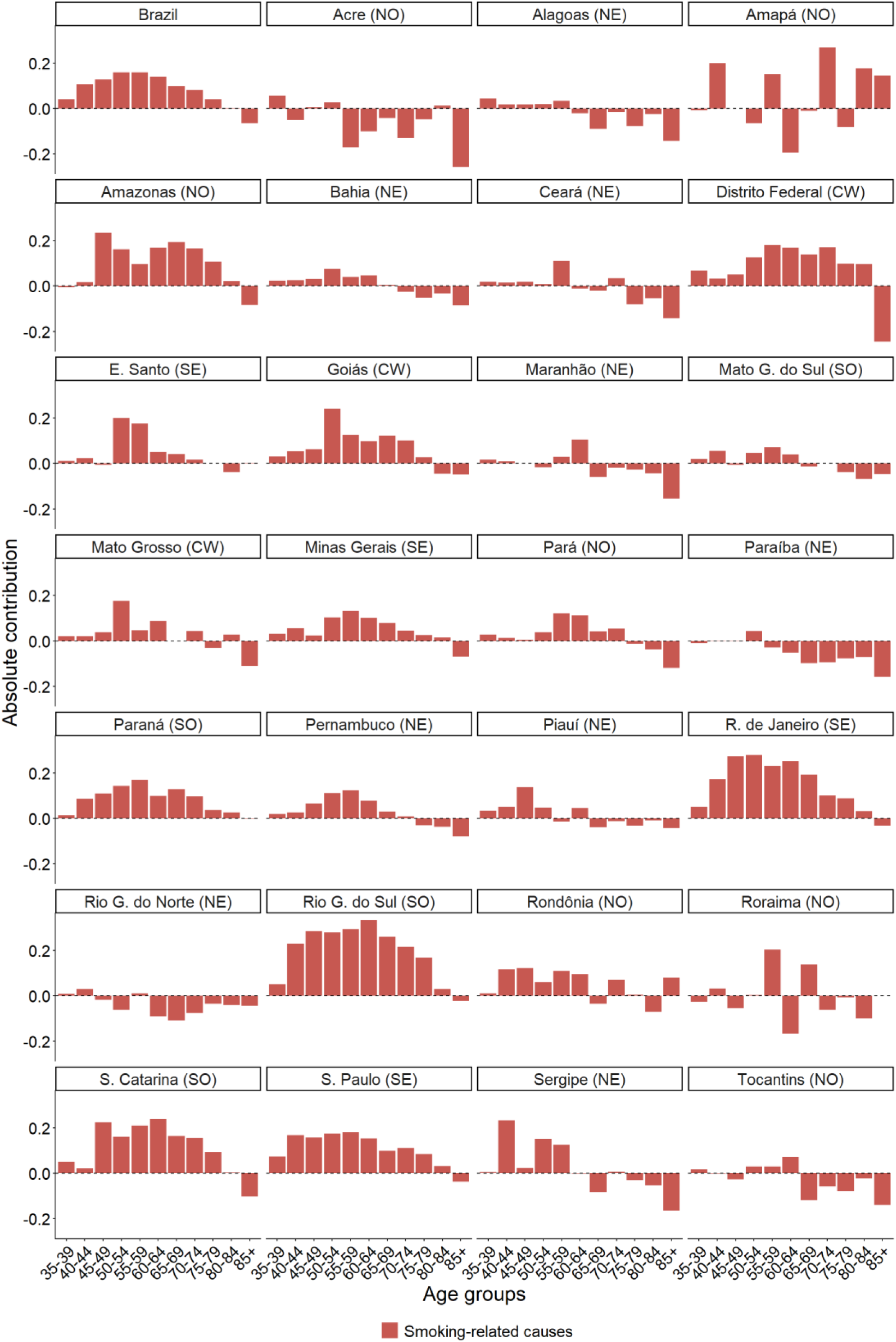
Contribution of smoking-related causes to the change in life expectancy at age 35 by age groups – Men, Brazilian states (2000–2023) Note: States are ordered alphabetically. NO = North, NE = Northeast, CW = Central-West, SE = Southeast, SO = South. Source: SIM/DATASUS/MS and IBGE.

**Figure D2.**
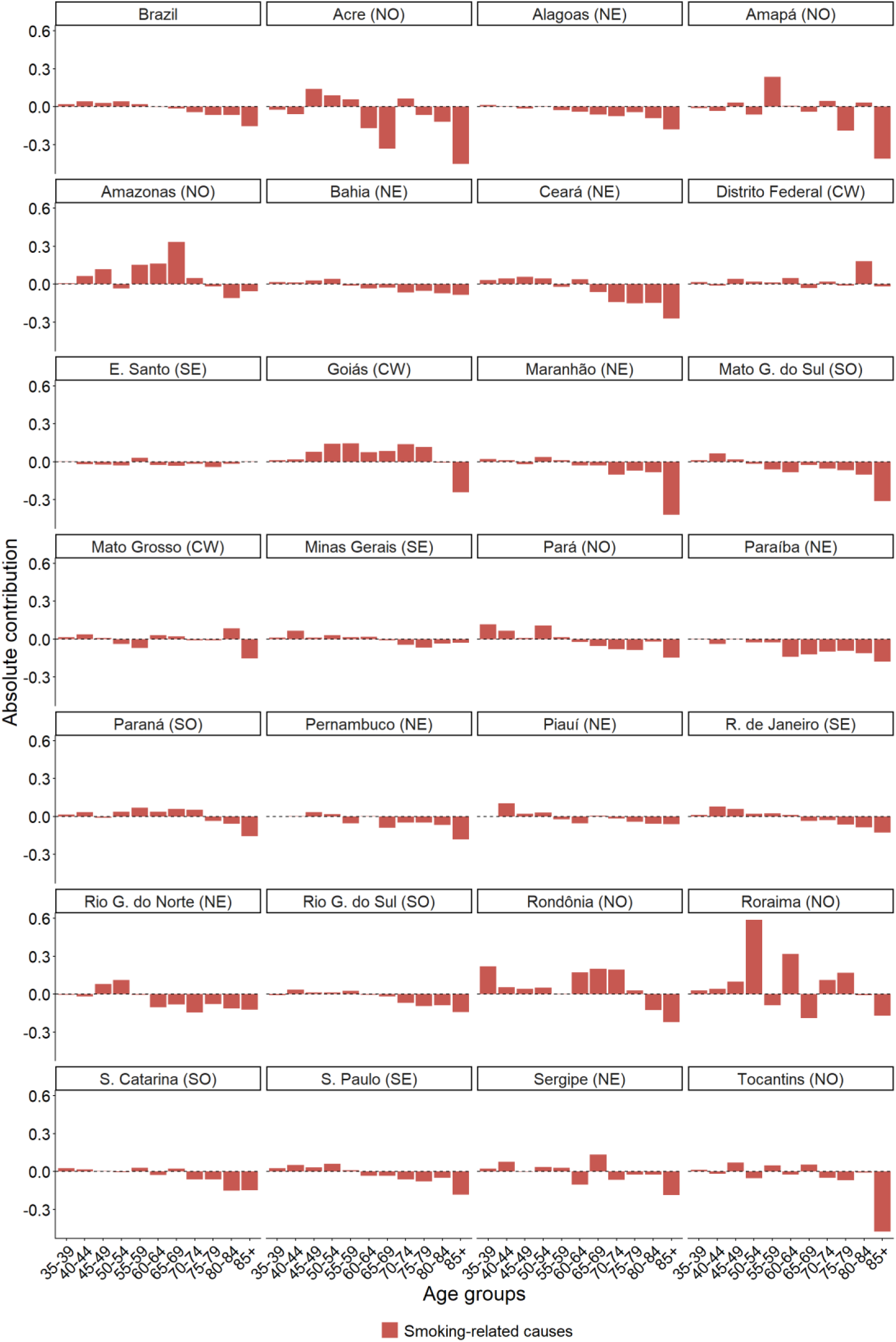
Contribution of smoking-related causes to the change in life expectancy at age 35 by age groups – Women, Brazilian states (2000–2023) Note: States are ordered alphabetically. NO = North, NE = Northeast, CW = Central-West, SE = Southeast, SO = South. Source: SIM/DATASUS/MS and IBGE.

## REFERENCES

1. Malta DC, Flor LS, Machado ÍE, et al. Trends in prevalence and mortality burden attributable to smoking, Brazil and federated units, 1990 and 2017. Population Health Metrics. 2020;18(suppl 1):24.

2. Malta DC, Gomes CS, Andrade FMDD, et al. Tabagismo no Brasil: percepções dos resultados de pesquisas domiciliares. REME Revista Mineira de Enfermagem. 2023;27:e-1518.

3. Maia EG, Stopa SR, de Oliveira Santos R, Claro RM. Trends in prevalence of cigarette smoking in Brazil: 2006-2019. American Journal of Public Health. 2021;111(4):730–738.

4. Dos Santos AMA, Triaca LM, Leivas PHS. How is smoking distributed in relation to socioeconomic status? Evidence from Brazil in the years 2013 and 2019. Economics & Human Biology. 2023;49:101240.

5. Lima FCSD, Silva DHND, Szklo AS, Scaff AJM, Reis RDS. Evolution of smoking and incidence of lung cancer in Brazil (2000-2020). Revista Brasileira de Cancerologia. 2025;71:e-114864.

6. World Health Organization. WHO global report on trends in prevalence of tobacco use 2000-2025. World Health Organization; 2020.

7. Reitsma MB, Fullman N, Ng M, et al. Smoking prevalence and attributable disease burden in 195 countries and territories, 1990-2015: a systematic analysis from the Global Burden of Disease Study 2015. The Lancet. 2017;389(10082):1885–1906.

8. Monteiro CA, Cavalcante TM, Moura EC, Claro RM, Szwarcwald CL. Population-based evidence of a strong decline in the prevalence of smokers in Brazil (1989-2003). Bulletin of the World Health Organization. 2007;85(7):527–534.

9. Portes LH, Machado CV, Turci SRB, Figueiredo VC, Cavalcante TM, Silva VLC. A Política de Controle do Tabaco no Brasil: um balanço de 30 anos. Ciência & Saúde Coletiva. 2018;23:1837–1848.

10. Wanderley-Flores B, Pérez-Ríos M, Montes A, et al. Mortalidad atribuida al consumo de tabaco en Brasil, 1996-2019. Gaceta Sanitaria. 2025;37:102297.

11. Lopez AD, Collishaw NE, Piha T. A descriptive model of the cigarette epidemic in developed countries. Tobacco Control. 1994;3(3):242–247.

12. Malta DC, Moura EC, Silva SA, Oliveira PPV. Prevalence of smoking among adults residing in the Federal District of Brasília and in the state capitals of Brazil, 2008. Jornal Brasileiro de Pneumologia. 2010;36(1):75–83.

13. Sao José BPD, Corrêa RDA, Malta DC, et al. Mortality and disability from tobacco-related diseases in Brazil, 1990 to 2015. Revista Brasileira de Epidemiologia. 2017;20(suppl 1):75–89.

14. Tam J, Jaffri MA, Mok Y, et al. Patterns of birth cohort-specific smoking histories in Brazil. American Journal of Preventive Medicine. 2023;64(4 suppl):S63-S71.

15. Peto R, Lopez AD, Boreham J, Thun M, Heath C Jr. Mortality from tobacco in developed countries: indirect estimation from national vital statistics. The Lancet. 1992;339(8804):1268–1278.

16. Reis CS. A história de tabagismo no Brasil segundo coortes de nascimento, sexo e escolaridade e seus efeitos prováveis sobre a mortalidade adulta futura [doctoral dissertation]. Universidade Federal de Minas Gerais; 2019.

17. Horiuchi S, Wilmoth JR, Pletcher SD. A decomposition method based on a model of continuous change. Demography. 2008;45(4):785–801.

18. Gaspar RS, Rezende LF, Laurindo FRM. Analysing the impact of modifiable risk factors on cardiovascular disease mortality in Brazil. PLOS ONE. 2022;17(6):e0269549.

19. Palloni A, Novak B, Pinto-Aguirre G. The enduring effects of smoking in Latin America. American Journal of Public Health. 2015;105(6):1246–1253.

20. Janssen F. Similarities and differences between sexes and countries in the mortality imprint of the smoking epidemic in 34 low-mortality countries, 1950–2014. Nicotine Tob Res. 2020;22(7):1210–1220.

21. Beltrán-Sánchez H, Finch CE, Crimmins EM. Twentieth century surge of excess adult male mortality. Proceedings of the National Academy of Sciences. 2015;112(29):8993–8998.

22. Janssen F. Changing contribution of smoking to the sex differences in life expectancy in Europe, 1950-2014. European Journal of Epidemiology. 2020;35(9):835–841.

23. Janssen F, El Gewily S, Bardoutsos A. Smoking epidemic in Europe in the 21st century. Tobacco Control. 2021;30(5):523–529.

24. Wanderlei-Flores B, Rey-Brandariz J, Corrêa PRP, et al. Smoking-attributable mortality by sex in the 27 Brazilian federal units: 2019. Public Health. 2024;229:24–32.

25. Sao José BPD, Corrêa RDA, Malta DC, et al. Mortality and disability from tobacco-related diseases in Brazil, 1990 to 2015. Revista Brasileira de Epidemiologia. 2017;20(suppl 1):75–89.

26. Borges GM. Health transition in Brazil: regional variations and divergence/convergence in mortality. Cadernos de Saúde Pública. 2017;33:e00080316.

27. Queiroz BL, Gonzaga MR, Vasconcelos AM, Lopes BT, Abreu DM. Comparative analysis of completeness of death registration, adult mortality and life expectancy at birth in Brazil at the subnational level. Population Health Metrics. 2020;18(suppl 1):11.

28. Mehta N, Elo I, Stenholm S, et al. International differences in the risk of death from smoking and obesity: the case of the United States and Finland. SSM-Population Health. 2017;3:141–152.

29. Preston SH, Glei DA, Wilmoth JR. A new method for estimating smoking-attributable mortality in high-income countries. International Journal of Epidemiology. 2010;39(2):430–438.

30. Morais ÉAHD, Oliveira BED, Roesberg JMA, et al. Fatores individuais e contextuais associados ao tabagismo em adultos jovens brasileiros. Ciência & Saúde Coletiva. 2022;27:2349–2362.

31. Espinoza-Derout J, Shao XM, Lao CJ, et al. Electronic cigarette use and the risk of cardiovascular diseases. Frontiers in Cardiovascular Medicine. 2022;9:879726.

32. Müller F, Wehbe L. Smoking and smoking cessation in Latin America: a review of the current situation and available treatments. International Journal of Chronic Obstructive Pulmonary Disease. 2008;3(2):285–293.

33. Giraldo-Osorio A, et al. Smoking-attributable mortality in South America: a systematic review. Journal of Global Health. 2021;11:04014.

## References

34. Ezzati, M., & Lopez, A. D. (2003). Estimates of global mortality attributable to smoking in 2000. The Lancet, 362(9387), 847–852.

35. Gorini, G., Chellini, E., Querci, A., & Seniori Costantini, A. (2003). Impatto dell’abitudine al fumo in Italia nel 1998: decessi e anni potenziali di vita persi. Epidemiol Prev, 27(5), 285–90.

36. Peto, R., et al. (1992). Mortality from tobacco in developed countries: indirect estimation from national vital statistics. The Lancet, 339(8804), 1268–1278.

37. Janssen, F., Trias-Llimos, S., & Kunst, A. E. (2021). The combined impact of smoking, obesity and alcohol on life-expectancy trends in Europe. International Journal of Epidemiology, 50(3), 931–941.

